# *GPATCH11* variants cause mis-splicing and early-onset retinal dystrophy with neurological impairment

**DOI:** 10.1101/2023.08.19.23293832

**Authors:** Andrea Zanetti, Lucas Fares-Taie, Jeanne Amiel, Jérôme Roger, Isabelle Audo, Matthieu Robert, Pierre David, Vincent Jung, Nicolas Goudin, Ida Chiara Guerrera, Stéphanie Moriceau, Danielle Amana, Nathalie Boddaert, Sylvain Briault, Ange-Line Bruel, Cyril Gitiaux, Karolina Kaminska, Nicole Philip-Sarles, Mathieu Quinodoz, Cristina Santos, Luisa Coutinho Santos, Sabine Sigaudy, Mariana Soeiro e Sá, Ana Berta Sousa, Christel Thauvin, Carlo Rivolta, Josseline Kaplan, Jean-Michel Rozet, Isabelle Perrault

## Abstract

Spliceosome and ciliary dysfunctions can lead to remarkably similar clinical syndromes. Studying ten individuals with retinal dystrophy, neurological involvement, and skeletal abnormalities, suggestive of both spliceosomopathies and ciliopathies, we involved GPATCH11, a lesser-known GPATCH-domain-containing regulators of RNA metabolism. To elucidate GPATCH11 function, we employed fibroblasts from unaffected individuals and patients carrying a recurring mutation specifically removing the main part of the GPATCH-domain while preserving other domains. Additionally, we generated a mouse model replicating the patient’s genetic defect, exhibiting behavioural abnormalities and retinal dystrophy. Our findings revealed GPATCH11 unique subcellular localization, marked as foci staining pattern and a diffuse presence in the nucleoplasm, alongside its centrosomal localization, indicating roles in RNA and cilia metabolism. We show dysregulation of U4 snRNA in patient cells and dysregulation in both gene expression and spliceosome activity within the mutant mouse retina, impacting key processes such as photoreceptor light responses, RNA regulation, and primary cilia-associated metabolism. These results highlight GPATCH11 roles in RNA metabolism, spliceosome regulation, and potential ciliary involvement. They underscore its significance in maintaining proper gene expression, contributing to retinal, neurological, and skeletal functions. Our research also demonstrates how studying rare genetic disorders can reveal broader gene functions, providing insights into GPATCH11 multifaceted roles.

## INTRODUCTION

The spliceosome is a large ribonucleoprotein complex that plays a crucial role in pre-mRNA splicing, intron excision, and exon ligation to generate functional mRNA. Comprising five core small nuclear ribonucleoprotein particles (snRNPs) and numerous other protein factors, called splicing factors, the spliceosome requires precise coordination for proper assembly, activation, and regulation to ensure accurate RNA splicing and cellular function^1^.

Spliceosomopathies refer to rare hereditary diseases that result from dysfunctions in the spliceosome. Despite the spliceosome being ubiquitous, spliceosomopathies tend to exhibit tissue-specific effects, notably impacting the survival of retinal rod photoreceptors, craniofacial development, or haematopoiesis^1^.

Among the spliceosome contributors, G-patch-domain-containing (GPATCH) proteins form a distinct group that is characterized by a glycine-rich motif and multiple RNA-binding motifs^2,3^. Despite sharing some common features, GPATCH proteins exhibit considerable variations in size, domain composition, and cellular localization^2^. Through their protein‒protein and protein-nucleic acid interactions, they contribute significantly to RNA metabolism by regulating the remodelling of RNAs and ribonucleoprotein complexes by the DEAH-box RNA helicases with which they associate^2,4–6^. In humans, 23 GPATCH-domain-containing proteins have been identified^2^. Of these, a dozen has been reported to be involved in pre-mRNA splicing and/or transcription regulation. Others participate in ribosome biogenesis, RNA export, rRNA and snoRNA maturation, and telomere maintenance^2,6,5^.

In line with the multifaceted functions of GPATCH proteins, several phenotypic traits have been liked to variations in GPATCH-coding genes, but only variants in *RBM10*^7^, *SON*^8^ have been definitively confirmed as disease-causing mutations in humans. Both of these genes have been linked to very severe multisystem developmental syndromes that may share similarities with spliceosomopathies but are notably more complex in their manifestations^7,8^.

GPATCH11, also known as coiled-coil-domain containing 75 (CCDC75) and centromere protein Y (CENP-Y), belongs to the less explored group of GPATCH-domain-containing proteins. While *GPATCH11* has been shown to be localized in the nucleus^9^, its specific role in RNA metabolism is not been demonstrated.

By analysis of whole exome sequencing (WES) in four affected families, with the objective of uncovering causal mutations, we found compelling evidence that recessive *GPATCH11* mutations are responsible for a syndrome characterized by early-onset and severe retinal degeneration, neurological issues, and abnormal craniofacial development.

To elucidate the function of *GPATCH11*, we used fibroblasts from patients who carry a mutation shared by three out of the four families. This mutation specifically removes part of the GPATCH-domain while preserving other domains. Additionally, we generated a mouse model replicating the patient’s genetic defect, exhibiting behavioural abnormalities and retinal dystrophy. Our findings revealed that GPATCH11 displays a unique subcellular localization, marked a foci staining pattern and a diffuse presence in the nucleoplasm, alongside with a centrosomal localization, indicating roles in spliceosome and cilia functions. Furthermore, we show dysregulation of U4 snRNA in patient cells and dysregulation in both gene expression and spliceosome activity within the mutant mouse retina, impacting key processes such as photoreceptor light responses, RNA regulation, and primary cilia-associated metabolism.

These results highlight GPATCH11 multifaceted roles in RNA metabolism, spliceosome regulation, and potential ciliary involvement. They underscore its significance in maintaining proper gene expression, contributing to retinal, neurological, and skeletal functions. Our research also demonstrates how studying rare genetic disorders can reveal broader gene functions, providing insights into GPATCH11 roles.

## RESULTS

### Biallelic *GPATCH11* mutations cause retinal dystrophy, neurodevelopmental delay, and behavioural problems with or without seizures and skeletal anomalies

The families were initially referred to a genetic consultation for distinctive presenting symptoms, namely, severe visual dysfunction from birth (Family 1), intellectual disability (Family 2), intellectual disability with encephalopathy (Family 3) and intellectual disability with retinitis pigmentosa (Family 4). However, a review of clinical files, re-examination and/or disease progression revealed common features of progressive retinal degeneration, intellectual disability, and dysmorphic features (**Fig.1**).

**Fig. 1:**
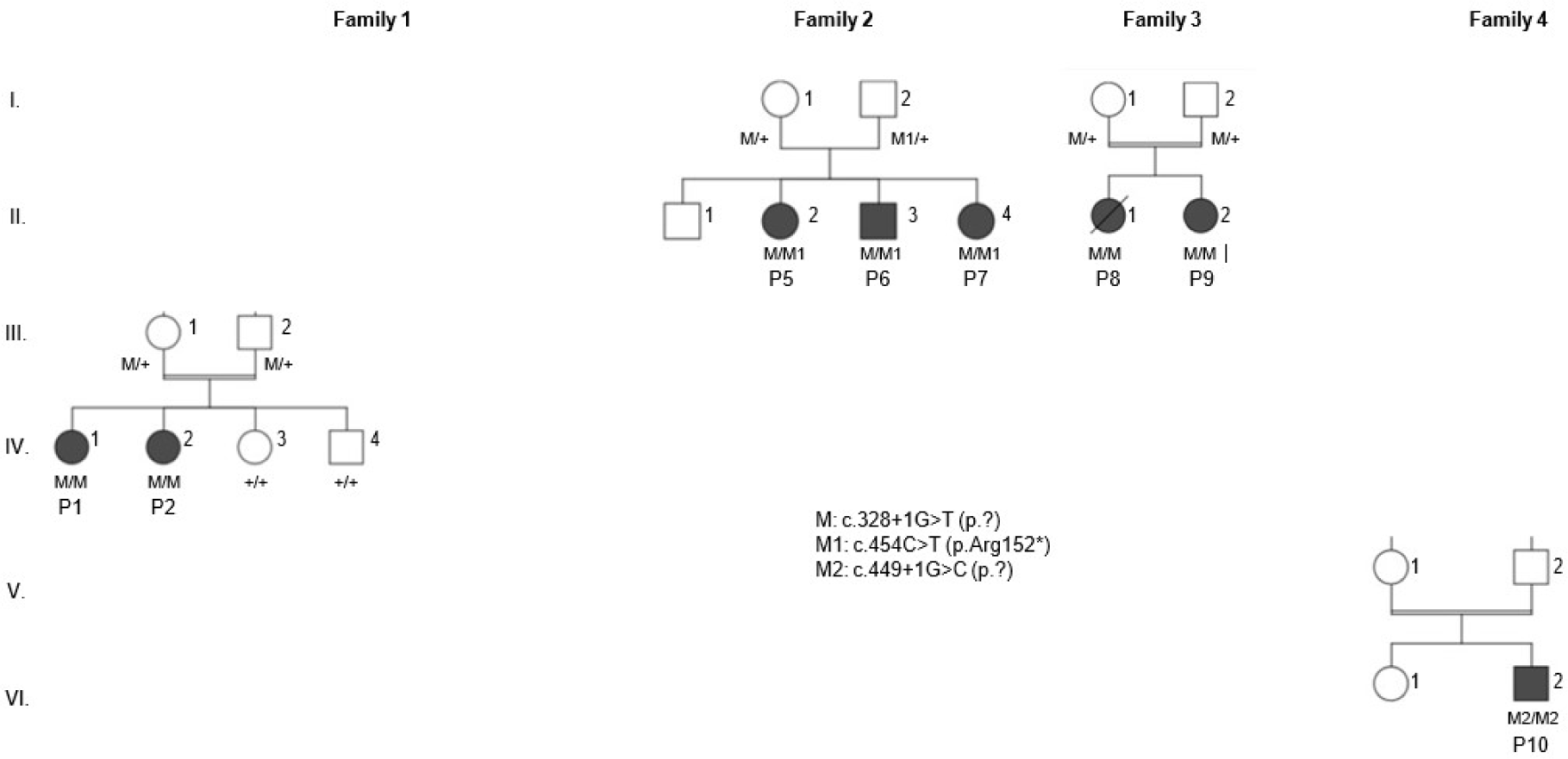
*GPATCH11* variants are responsible for syndromic inherited retinal diseases. Pedigree and segregation analysis of the four families carrying *GPATCH11* variants. M: c.328+1G>T (p.?); M1: c.454C>T (p.Arg152*); M2: c.449+1G>C (p.?); +, wild-type allele; P, patient. F1:IV-1 (P1) and F1:IV-2 (P2) are affected individuals whose fibroblasts were analyzed.

The index case in Family 1 (F1:IV-2) and an elder affected sibling (F1:IV-1) manifested severe visual deficiency with nystagmus and altered electroretinography (ERG) results in the first months of life (**Table 1**). Ophthalmological evaluation in childhood revealed poor vision (visual acuity of 1/20 and hand movements during their childhood, for F1:IV-1 and F1:IV-2 respectively; **Table 1**), macular atrophy and peripheral pigmentary deposits at the fundus (**Fig. 2a**), and outer retinal thinning affecting predominantly the perimacular region at SD-OCT (**Fig. 2b**). As they grew up, the two affected individuals manifested agitation, hyperactivity, frustration, and delay in language acquisition (**Table 1**). Neuro-pediatric workup and neuroimaging revealed neurodevelopmental anomalies with normal cerebral MRI. Recently, the elder affected child manifested seizures. Their affected paternal relatives (F1:III-3 and F1:III-6; **Fig. 1**), who were born to a first-cousin marriage, were seen in a north African hospital for severe visual dysfunction and marked neurodevelopmental delay manifesting as poor language skills, mild ataxia and general movement disorganization. The eldest affected relative passed away in the second decade of life during an episode of fever (**Table 1**).

**Table 1:** Summary of clinical features of ten affected individuals from four families with *GPATCH11* variants. Ophthalmological, neurological and dysmorphological investigation in all affected individuals. Abbreviation used: NA, not available; NP, not performed.

| Individual | F1:III-3 | F1:III-6 | F1:IV-1 | F1:IV-2 | F2:II-2 | F2:II-3 | F2:II-4 | F3:II-1 | F3:II-2 | F4:II-2 |
| --- | --- | --- | --- | --- | --- | --- | --- | --- | --- | --- |
| Age range | Dead in the second decade of life because of fever | In the second decade of life | In the second decade of life | In the first decade of life | In the first decade of life | In the first decade of life | In the first decade of life | Dead in the third decade of life because of stroke | In the third decade of life | In the fourth decade of life |
| **Ophthalmological Features** |  |  |  |  |  |  |  |  |  |  |
| Age at diagnosis | N.A. | Birth | First months of life | Birth | First years of life | Birth | Birth | N.A. | N.P. | Second decade of life |
| Nystagmus | N.A. | + | + | + | N.A. | N.A. | N.A. | N.A. | N.A. | N.A. |
| Photophobia | N.A. | N.A. | + | N.A. | N.A. | N.A. | N.A. | N.A. | N.A. | + |
| Night blindness | N.A. | N.A. | N.A. | N.A. | N.A. | + | N.A. | N.A. | N.A. | + since early childhood; color vision trouble since the second decade of life |
| Visual acuity (RE) (LE) | N.A. | N.A. | 1/20 | Hand movements | 4/10; hypermetropia | 8/10 | N.A. | Myopia | Myopia | 0,4 OD; 0,1 OS |
| Refractive error | N.A. | N.A. | +8.75 (-1.5) 5°<br>+8.5 (-1.25) 170° | +1.25 (-0.25) 45°<br>+0.00 (-0.75) 170° | +1 (-0.50) 140°<br>-0.50 (-0.75) 75° | +1<br>+1.25 | N.A. | N.A. | N.A. | N.A. |
| ERG electronegativity | N.P. | N.A. | Flat | Flat | Flat | N.A. | N.A. | N.A. | N.A. | N.A. |
| Fundus | N.A. | N.A. | Macular atrophy; pigmentary deposits in periphery | Macular atrophy; pigmentary deposits in periphery | Pigmentary deposits in periphery; light foveolar hypoplasia | N.A. | N.A. | N.A. | Optic atrophy; pigmentary deposits; bruised vessels | Retinitis pigmentosa signs: moderately pale optic nerve, narrowed retinal vessels, peripheral pigmentation (bone spicules) |
| Macular SD-OCT | N.P. | N.P. | Global alteration of the retina | N.P. | N.A. | N.A. | N.A. | N.A. | N.A. | Decreased retinal thickness (mainly external layers with relative foveal sparing OD) |
| Autofluorescence | N.A. | N.A. | Hyper autofluorescence of the macula surrounded by hypo autofluorescence | Hyper autofluorescence of the macula surrounded by hypo autofluorescence | Hyper autofluorescence of the macula | N.A. | N.A. | N.A. | N.A. | N.A. |
| **Neurological features** |  |  |  |  |  |  |  |  |  |  |
| Neurological examination | Psychomotor retardation; intellectual disability; no language | Psychomotor retardation; intellectual disability; no language | IME Psychomotor retardation; language disorder; | IME Psychomotor retardation; language disorder | ULIS Psychomotor retardation; intellectual disability; | Language: do sentences | N.A. | Psychomotor retardation; discrete ataxia; encephalopathy | Psychomotor retardation; ataxia; encephalopathy; autism | Cognitive deficit, global developmental delay; mainstream |
|  |  |  | hyperactivity;<br>epilepsy:<br>continuous<br>spikes and<br>waves during<br>sleep |  | language<br>disorder |  |  | hy;<br>retardation;<br>language:<br>says few<br>words;<br>epilepsy at<br>few months<br>old | spectrum<br>disorder; self-<br>and hetero-<br>aggression;<br>retardation;<br>no language;<br>epilepsy | schooling<br>until the<br>second<br>decade of life;<br>currently<br>autonomous<br>for daily<br>activities. |
| MRI | N.A. | N.A. | Normal | Normal | Normal | Normal | N.A. | Hypoplasia of<br>frontal lobes | N.A. | Normal |
| **Dysmorphological features** |  |  |  |  |  |  |  |  |  |  |
| Morphological<br>examination | Facial<br>dysmorphia | Facial<br>dysmorphia | Facial<br>dysmorphia;<br>enophtalmia;<br>short<br>philtrum;<br>large incisors;<br>diastema;<br>oligodontia;<br>flat feet;<br>hyperlaxity | Facial<br>dysmorphia;<br>enophtalmia;<br>short<br>philtrum;<br>large incisors;<br>diastema;<br>enamel<br>dysplasia; flat<br>feet;<br>hyperlaxity | Facial<br>dysmorphia;<br>enophtalmia;<br>short<br>philtrum;<br>large incisors;<br>broad ovula;<br>flat feet | Facial<br>dysmorphia | N.A. | Facial<br>dysmorphia;<br>enophtalmia;<br>thick lips;<br>prominent<br>columella;<br>short<br>philtrum;<br>large incisors;<br>hyperlaxity;<br>short stature:<br>143 cm | Facial<br>dysmorphia;<br>enophtalmia;<br>thick lips;<br>prominent<br>columella;<br>short<br>philtrum;<br>large incisors;<br>abdominal<br>obesity; short<br>stature: 146<br>cm | Some<br>dysmorphic<br>features<br>noticed (not<br>suggestive of<br>a specific<br>syndrome),<br>macrocephaly<br>, large<br>external ears,<br>small pre-<br>auricular<br>tubercle on<br>the right side,<br>small and<br>deep-set eyes,<br>long and<br>prominent<br>nose with<br>long<br>columella,<br>hypoplastic<br>nasal alae,<br>short 5th toes |
| **Other** |  |  |  |  |  |  |  |  |  |  |
| Metabolism<br>examination | N.A. | N.A. | N.A. | N.A. | N.A. | N.A. | N.A. | Diabetes<br>discovered in<br>the second<br>decade of life | Diabetes<br>discovered in<br>the second<br>decade of life | N.A. |

**Fig. 2:**
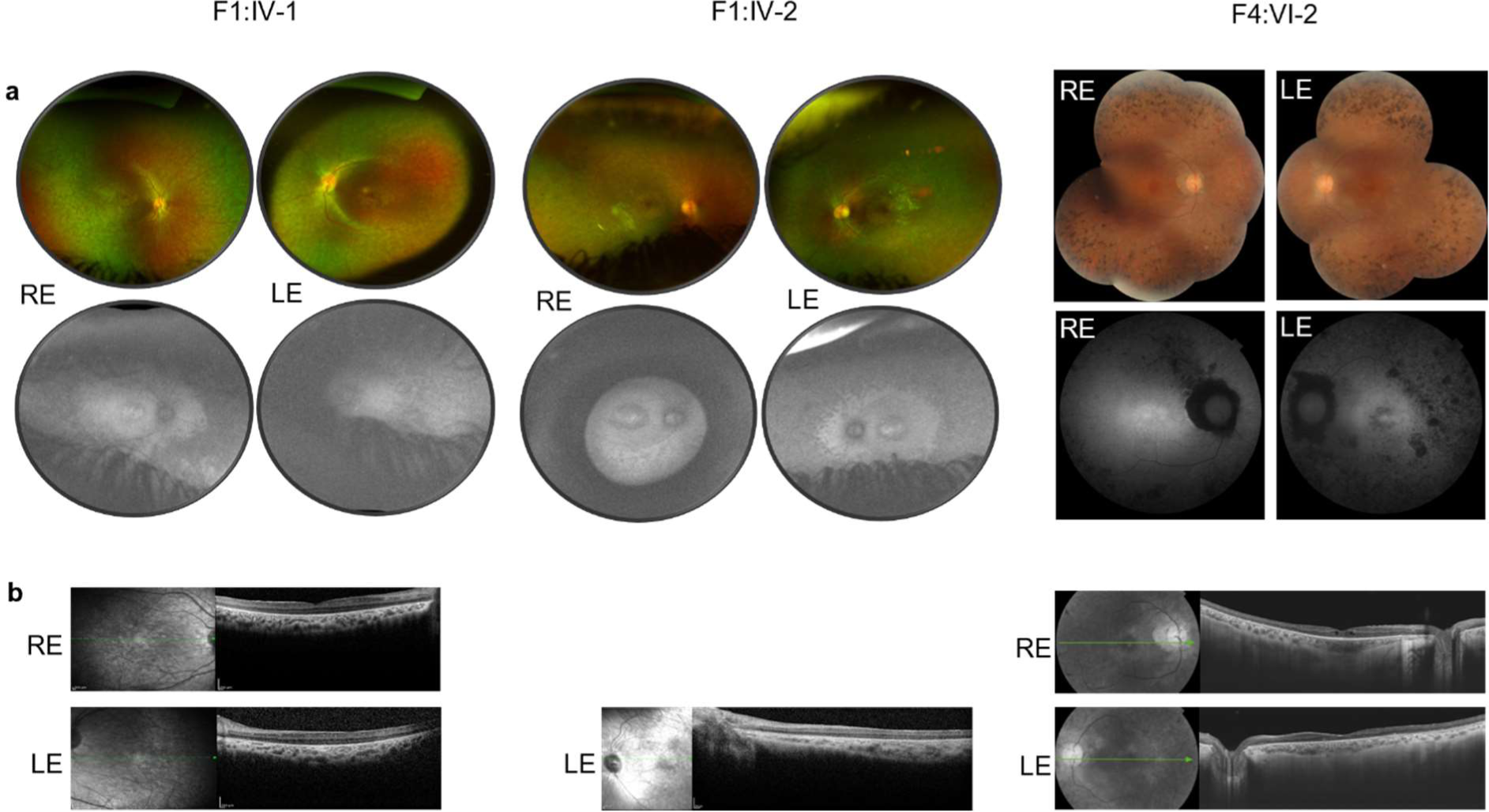
Clinical analysis of affected individuals. (**a**) Color fundus photographs of the affected individuals F1:IV-1 (P1), F1:IV-2 (P2) and F4:VI-2 (P10). RE: right eye; LE: left eye. (**b**) Spectral-domain optical coherence tomography (SD-OCT) of the macular region of the retina of the affected individuals F1:IV-1 (P1), F1:IV-2 (P2) and F4:VI-2 (P10). (**c**) Clinical images of affected individuals F1:IV-1 (P1), F1:IV-2 (P2) and F3:II-2 (P9) carrying the homozygous c.328+1G>T (p.?) *GPATCH11* variant and of affected individual F2:II-2 (P5) carrying the variant c.328+1G>T (p.?) in *trans* with c.454C>T (p.Arg152*).

The three affected siblings of Family 2 (**Fig. 1**) were initially addressed for intellectual disability, speech and walking delay and dyspraxia. The eldest affected subject (F2:II-2) was reported to have displayed normal electroencephalography (EEG) and MRI. Before 5 years old, she manifested visual problems, and ophthalmological examination revealed reduced visual acuity (4/10 left and right eyes, LRE), hypermetropia and flat ERG (**Table 1**). The eldest of the two affected siblings in Family 3 (F3:II-1; **Fig. 1**) presented with short stature, psychomotor retardation, encephalopathy and seizures, lack of muscular coordination for voluntary movement in early childhood, and diabetes in the second decade of life, and she died in the third decade of life of thalamic stroke. The younger affected chld (F3:II-2) presented the same clinical features (**Table 1**). She and the affected subjects from Families 1 (F1:IV-1, F1:IV-2) and 2 (F2:II-2) were examined by dysmorphologists who noted similar facial dysmorphic features consisting of enophthalmos, a short philtrum, large incisors and diastema. The affected children in Family 4 (F4:VI-2, **Fig. 1**) were reported to display night blindness since childhood but were diagnosed of retinitis pigmentosa (RP) with peripheral deposit of pigments in the second decade of life (**Fig. 2a-b**). The cognitive deficits with global developmental delay were noticed in the first years of life and dysmorphic features were reported (**Table 1**).

Analysis of exome datasets from affected individuals and their unaffected relatives identified the *GPATCH11* splice-site variant c.328+1G>T (p.?) (NM_174931.4) in homozygosity and in compound heterozygosity with the nonsense variant c.454C>T (p.Arg152*) in affected subjects from consanguineous Families 1 and 3 and nonconsanguineous Family 2, respectively (**Fig. 1**). Haplotype reconstruction using exome datasets from the three families, all of whom originate from Maghreb, identified a short (1.27 Mb) common haplotype encompassing the splice-site variant c.328+1G>T (**Supplementary Table 1**), supporting an ancient founder effect. In Family 4, originating from Europe, exome analysis identified homozygosity for another splice-site variant, c.449+1G>C (p.?) (**Fig. 1**). Consistent with autosomal recessive disease transmission, the parents of affected individuals in the four families carried these variants in single heterozygosity (**Fig. 1**).

In the index patient from the Family 4, originating from Europe, analysis of WES data identified homozygosity for another splice-site variant, c.449+1G>C, (p.?) in *GPATCH11* (**Fig. 1**). Consistent with autosomal recessive disease transmission, the parents of affected individuals in the four families carried these variants in single heterozygosity (**Fig. 1**). Whole exome sequencing in four independent families allowed the identification of 3 different novel variants in *GPATCH11*.

The three variants were absent from controls in the genetic variation datasets (Exome Sequencing Project, 1000 Genomes Project, and Exome Aggregation Consortium) tested by Alamut Visual™. *In silico* predictions suggest that the consensus donor splice-site variants c.328+1G>T (Donor loss: 0,95; SpliceAI) and c.449+1G>C (Donor loss: 1; SpliceAI) cause skipping of in-frame exon 4 and out-of-frame exon 5, respectively.

### Cells from individuals carrying the c.328+1G>T variant produce a mutant GPATCH11 protein lacking the internal portion of the GPATCH-domain

To validate the *in silico* predictions described above, we performed RT-PCR analysis of RNA isolated from the fibroblasts of the affected siblings in Family 1 F1:IV-1 (P1) and F1:IV-2 (P2), carrying the c.328+1G>T variant in homozygosity. This analysis showed skipping of the 42 bp-long exon 4, encoding a 14 amino acid-long sequence (aa 96–109) that links the helix and loop braces of the GPATCH-domain encoded by the adjacent exons^2^ (**Fig. 3a, c**).

**Fig. 3:**
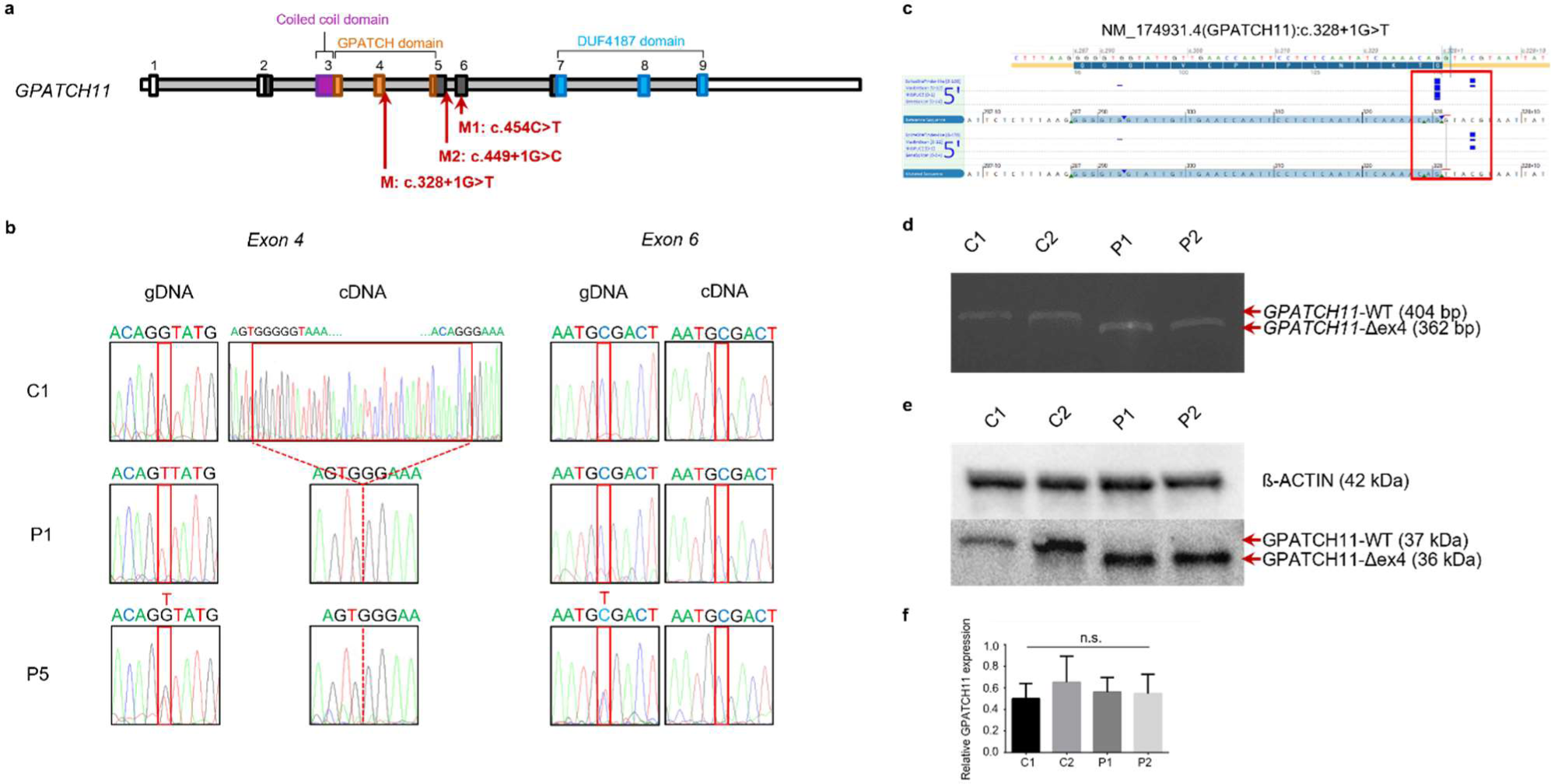
Presentation of *GPATCH11* variants and molecular analysis of the c.328+1G>T splice-site change. (**a**) Human *GPATCH11* gene diagram indicating the positions of c.328+1G>T, c.454C>T, and c.449+1G>C variants relative to NM 174931.4 transcript. (**b**) Representative chromatogram of gDNA and cDNA sequences from exon 4 and exon 6 of one control (C1) and affected individuals carrying the c.328+1G> T (exon 4) in homozygosity (P1) and in compound heterozygosity with the c.454C>T (exon 6) (P5) variants. (**c**) Electrophoresis of *GPATCH11* cDNA showing detection of a shorter mRNA (*GPATCH11*-Δex4 (362 bp)) in patients’ (P1, P2) fibroblasts as compared to the control (*GPATCH11*-WT (404 bp)) in controls (C1, C2). (**d**) Western Blot analysis of protein extracts using GPATCH11 antibody targeting residues 111-192 showing detection of shorter protein isoform (GPATCH11-Δex4 (36 KDa)) in patients’ fibroblasts (P1, P2) as compared to the controls (GPATCH11-WT (37 KDa)) in controls (C1, C2). (**e**) Abundance of GPATCH11 products in affected individual fibroblasts is like controls. Bars represent mean±SEM from three experimental replicates. n.s., not significant.

Western blot analysis using an antibody specific to a sequence downstream of the GPATCH-domain detected a unique band in patient cells. The molecular weight corresponding to this band was lower than that of the wild-type counterpart in control fibroblasts and consistent with a 14-residue deletion (**Fig. 3d**). The abundance of the mutant protein in fibroblasts from the affected individuals F1:IV-1 (P1) and F1:IV-2 (P2) and controls were not significantlly different according to Western Blot analysis (**Fig. 3e**).

Analysis of blood RNA from the affected subject of Family 2 (F2:II-2; P5) showed apparent homozygosity for the variant c.328+1G>T, suggesting nonsense-mediated mRNA decay (NMD) of the mRNA transcribed for the trans allele carrying the c.454C>T (p.Arg152*) nonsense variant (**Fig. 3b**).

Overall, our data show that the variant c.328+1G>T produces a mutant (shorter) GPATCH11 protein, whereas the variant c.454C>T likely triggers NMD and therefore results in no GPATCH11 protein.

### GPATCH11 displays a distinctive subcellular localization characterized by a foci staining pattern and centriolar localization, resilient to ablation of GPATCH- or CCDC-domains

To analyse the expression of GPATCH11 in cellular models, we conducted confocal microscopy analysis of immunostained control fibroblasts, using both methanol and PFA fixation methods. This examination not only confirmed the expected nuclear localization of the GPATCH11 protein^9^ but also revealed a distinctive foci staining pattern (through methanol fixation) and a diffuse presence in the nucleoplasm, in addition to a centrosomal localization. This distribution pattern aligns with what has been previously described for centrosome-associated spliceosome components^10,11^. Of note, basal body staining was decreased in PFA-fixed cells (**Supplementary Fig. 1a**). To refine the nucleoplasm localization of the protein, fibroblasts were immunostained for GPATCH11 together with SC-35 (speckles), H3K9me3 (heterochromatin) and γH2AX (DNA damage) antibodies (**Supplementary Fig. 1b**). GPATCH11 is detected closed to speckles and heterochromatin. To refine the subcellular localization of the protein, fibroblasts were immunostained for GPATCH11 together with the cilium axoneme, centrosome and centrosome linker using ACETYLATED-α-TUBULIN, CENTRIN-3 and ROOTLETIN antibodies, respectively. Stimulated-emission-depletion (STED) microscopy detected GPATCH11 in the centrosome linker region anchoring numerous coiled-coil proteins to the proximal end of centrioles, including notable localization closed to ROOTLETIN (**Fig. 4**).

**Fig. 4:**
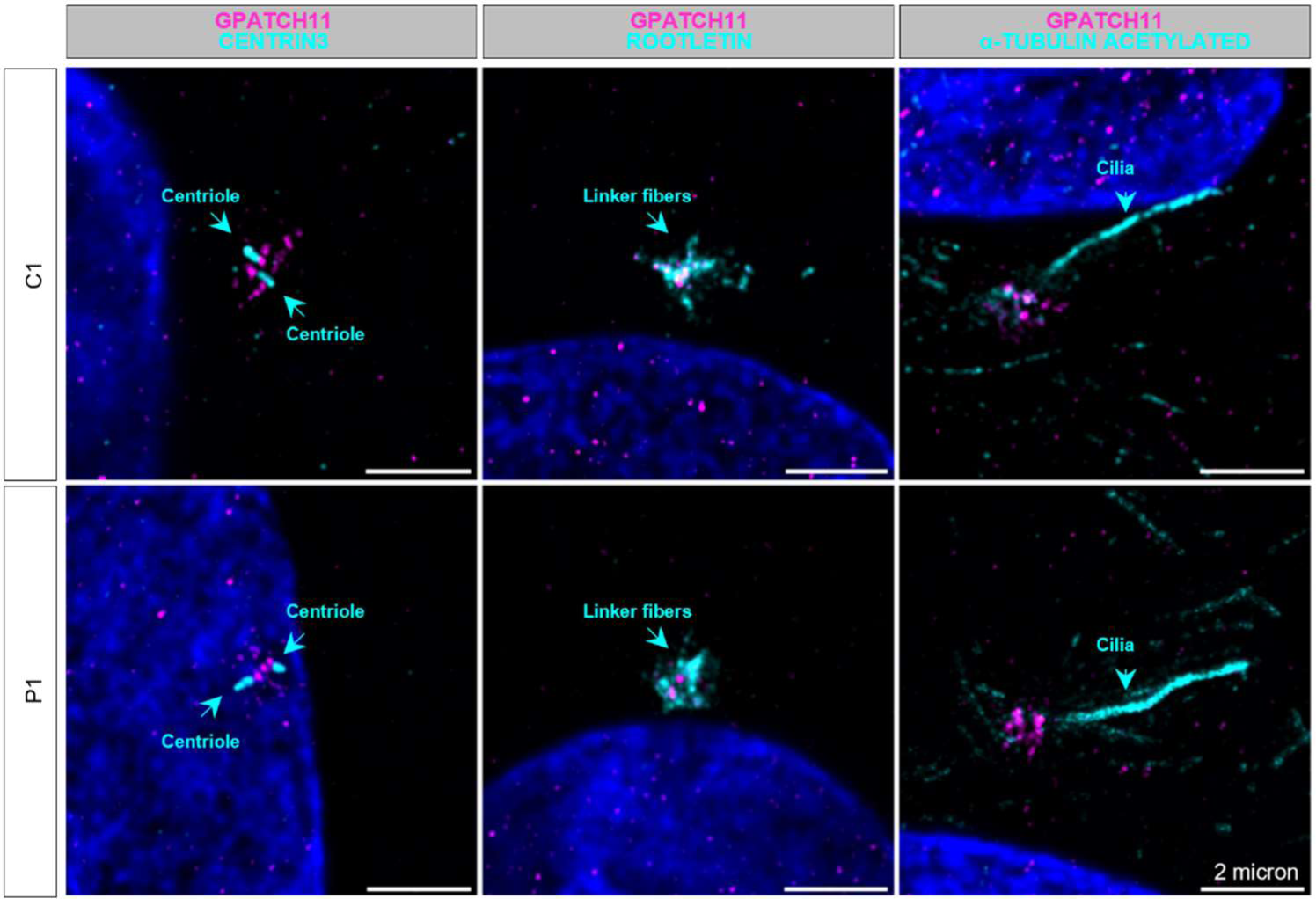
Centrosomal localization of GPATCH11. Immunostaining of GPATCH11 (magenta), CENTRIN3, ROOTLETIN and ACETYLATED α-TUBULIN (cyan) proteins in the fibroblasts from a control (C1) and affected individual carrying the c.328+1G>T variant in homozygosity (P1) following 24 hours of serum-free culture to promote ciliation. DAPI is used to label the nucleus. Scale bars, 2µm

To determine whether the GPATCH-domain has a role in the localization of the protein, the fibroblasts from the siblings F1:IV-1 (P1) and F1:IV-2 (P2) carrying the c.328+1G>T mutation in homozygosity were immunostained using the GPATCH11 antibody. The subcellular distribution of the protein and its abundance, as determined using a machine learning with Ilastik^12^ were comparable to control fibroblasts (**Fig. 4; Supplementary Fig. 1a, c**).

Given that many coiled-coil (CCDC) domain-containing proteins have shown centrosomal localization^13,14^, we conducted an assessment to determine whether the CCDC-domain of GPATCH11 is involved in its basal body localization. In this aim, we generated a homozygous in-frame deletion of the coding sequence in exon 3 was generated by the CRISPR‒Cas9 technique in the hTERT-RPE1 cell line and confirmed the presence of shortened mRNA and protein was confirmed by RT‒PCR and Western blot analysis, respectively (**Supplementary Fig. 2a, b**). Analysis of the subcellular localization of the Δ-CCDC GPATCH11 isoform in these cells revealed an unaltered distribution as compared to the wild-type counterpart in control hTERT-RPE1 cells (**Supplementary Fig. 2c**).

Collectively, these findings suggest that GPATCH11 is a spliceosome component associated with the centrosome, and that neither the GPATCH-nor the CCDC-domain plays a role in its subcellular localization.

### Cilia biogenesis and function are not impacted by the alteration of the GPATCH- or CCDC-domains

Centrosome-associated spliceosome components have previously been reported as regulators of ciliogenesis^11^. Therefore, we aimed to investigate whether the GPATCH- or CCDC-domains of GPATCH11 play a role in cilia formation. To achieve this, fibroblasts from homozygous F1:IV-1 (P1) and F1:IV-2 (P2) individuals, as well as Δ-CCDC hTERT-RPE1 cells, were cultured in a serum-free medium to promote cilia formation. Immunostaining was performed using PERICENTRIN and ACETYLATED Α-TUBULIN antibodies to label the primary cilium basal body and axoneme, respectively. The analysis revealed that the abundance of ciliated cells and the mean axonemal length were not significantly different from those detected in controls. This suggests that neither alterations in the GPATCH-domain nor deletion of the CCDC-domain had an impact on ciliation (**Supplementary Fig. 2d, e; Supplementary Fig. 3a, b**).

The Sonic Hedgehog (SHH) pathway is a critical developmental signalling pathway functionally linked to primary cilia in vertebrates^15^. To assess whether alterations in the GPATCH- or CCDC-domains had any impact on the functionality of the SHH signalling pathway in relation to primary cilia, F1:IV-1 (P1), F1:IV-2 (P2) and Δ-CCDC hTERT-RPE1 cells were treated using smoothened (SM) agonist known as SAG, which is a potent SHH activator. Following treatment, we measured the transcription levels of key genes involved in the SHH pathway, including *GLI1*, *GLI2*, *SMO*, and *PTCH1*, using RT-qPCR. The results of our quantitative gene expression analysis revealed that, despite alterations in the GPATCH- or CCDC-domains, the SHH pathway showed normal activation in patient fibroblasts when compared to control counterparts (**Supplementary Fig. 4**). This finding suggests that the SHH pathway retains its functionality in the presence of these domain alterations.

Collectively, this analysis suggests that neither the structure nor the function of primary cilium is impacted by alterations in the GPATCH- or CCDC-domains.

### Expression of the U4 snRNA is impacted by the disruption of the GPATCH-domain in patients’ cells

To assess whether the loss of the central part of the GPATCH-domain had an impact on the expression of small nuclear RNAs (snRNAs) associated with the spliceosome, we quantified U1, U2, U4, U5, and U6 snRNAs in cells from F1:IV-1 (P1) and F1:IV-2 (P2) individuals, as well as controls, using RT-qPCR with specific primers. Our analysis revealed a statistically significant reduction in the abundance of U4 snRNA and a trend toward reduced levels of U5 snRNA in cells from the two patients compared to controls. In contrast, the abundance of U1, U2, and U6 snRNAs remained similar between patient and control fibroblasts (**Fig. 5**). These findings provide further support for the role of GPATCH11 in the spliceosome.

**Fig. 5:**
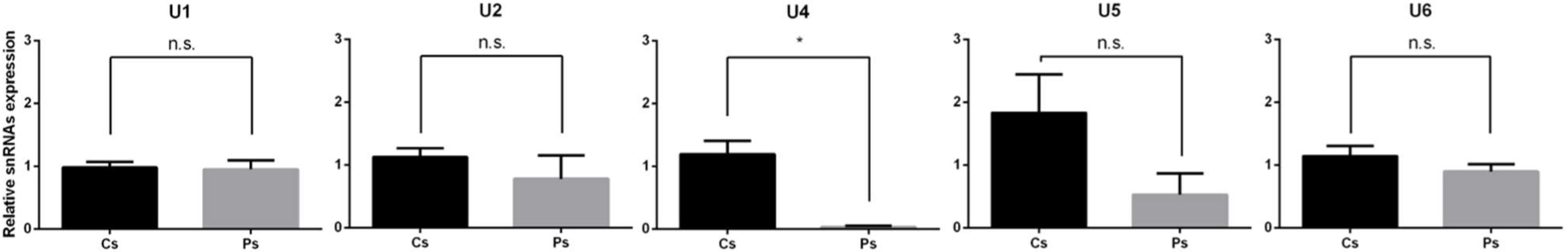
snRNAs analysis. RT-qPCR analysis of *U1*, *U2*, *U4, U5* and *U6* mRNAs abundance in fibroblasts from four controls (Cs) and P1 and P2 affected individuals (Ps). Graphic bars show the mean±SEM from three independent experiments. n.s., not significant. * p≤0.05.

### Mice expressing a GPATCH11 protein analogous to the human mutant variant experience rapid retinal degeneration, memory impairment, and infertility

As described previously, the c.328+1G>T variant is responsible for the skipping of exon 4 in humans. To model this variant *in vivo*, the corresponding exon in the mouse genome (exon 5) was ablated by the Cas9-mediated double-cut exon deletion technique using intronic guides located on either side of the target exon (**Fig. 6a**). Mice carrying exon 5 deletion in homozygosity (*Gpatch11^Δ^*^5^*^/Δ^*^5^) were viable and developed normally (the aspect, the weight and their daily behaviour were normal). However, males were completely infertile and exhibited smaller than normal and empty testes (**Supplementary Fig. 5**). In contrast, females exhibited normal fertility.

**Fig. 6:**
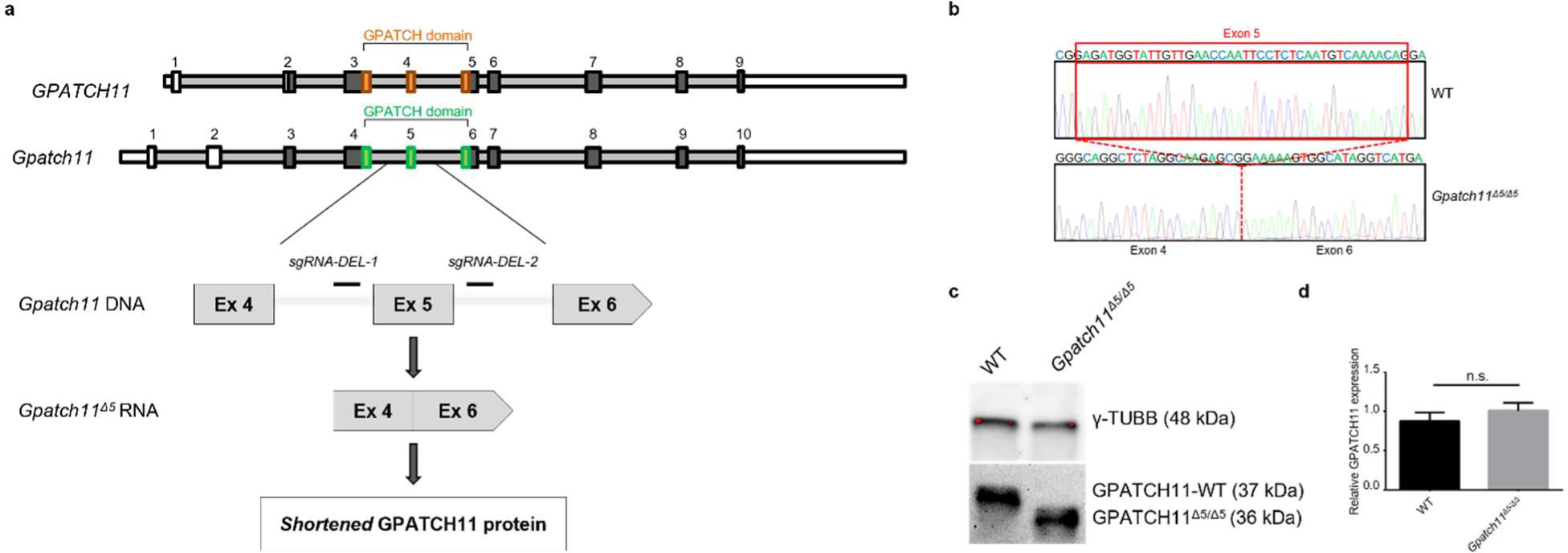
Generation of a mouse model expressing a protein analogous to the human mutant variant. (**a**) Diagram depicting the human *GPATCH11* gene and the murine *Gpatch11* gene. The schematic illustrates CRISPR/Cas9-mediated exon 5 deletion from the mouse Gpatch11 locus (human exon 4 corresponds to murine exon 5). (**b**) Representative chromatogram displaying wildtype (WT) and homozygous mutant (*Gpatch11^Δ^*^5^*^/Δ^*^5^) mice cDNA sequences amplified from retina samples using forward and reverse primers in exon 3 and 8, respectively. (**c**) Detection and (**d**) quantification of GPATCH11 wildtype and homozygous mutant isoforms relative to γ-TUBULIN4B by Western blot analysis of retina protein extracts. Bars indicate mean±SEM calculated from three experimental replicates. n.s., not significant.

The expected exclusion of exon 5 from the mRNA and presence of a shortened protein product in the retina of *Gpatch11^Δ^*^5^*^/Δ^*^5^ mice were verified by RT‒PCR and Sanger sequencing and Western blot analysis, respectively (**Fig. 6b, c**). The abundance of the mutant protein in *Gpatch11^Δ^*^5^*^/Δ^*^5^ mouse retina and of the wild-type counterpart in the retinas of the wild-type control littermates were not significantly different according to Western blot analyses (**Fig. 6c, d**).

Considering the retinal defects found in human patients, we characterized the retinal phenotype of *Gpatch11^Δ^*^5^*^/Δ^*^5^ mice through longitudinal functional and structural studies based on electroretinography (ERG), histology and immunohistochemistry analysis using wild-type littermates as controls. *Gpatch11^Δ^*^5^*^/Δ^*^5^ pups displayed normally developed and layered retinas at the age of 15 days (1 day from eye opening), with normal rod-specific but moderately reduced cone-specific responses to light stimulation (**Fig. 7a, b**). Rod and cone responses declined progressively as the mice aged, the photoreceptor nuclear layer became slimmer, and the RHODOPSIN and S and M cone OPSIN immunostaining weakened, supporting the gradual loss of rod and cone photoreceptors, respectively. By the age of 3 months, approximately half of the photoreceptor nuclei were lost, and the depletion of the opsin content was even more severe in rods and, more importantly, in cone outer segments, where light is captured and transduced (**Supplementary Fig. 6**). Consistently, ERG responses were highly altered. At the age of 6 months, ERG responses were flat, and photoreceptor nuclei were almost completely lost (**Fig. 7a, b**).

**Fig. 7:**
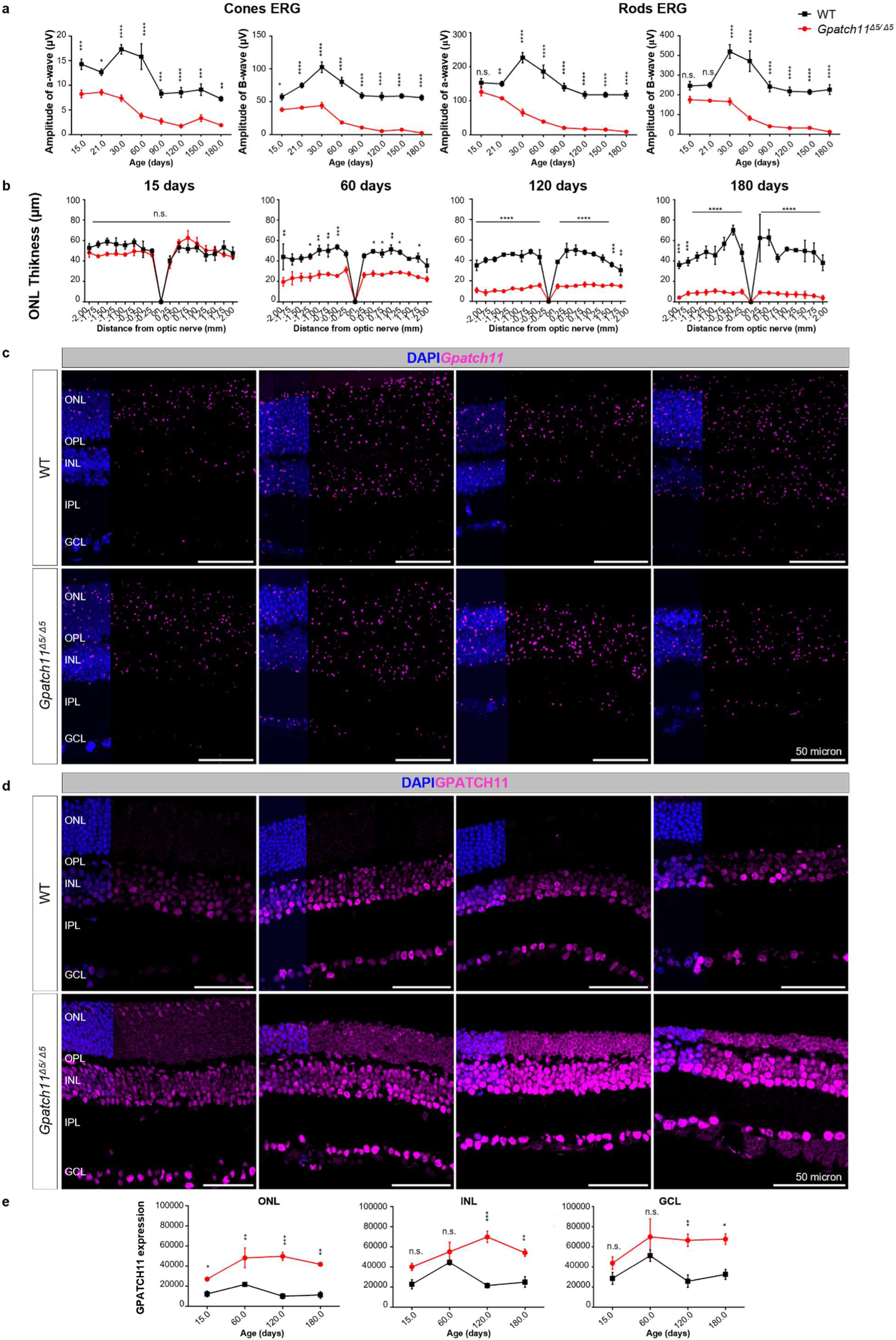
Longitudinal analysis of retinal structure and function in *Gpatch11^Δ^*^5^*^/Δ^*^5^ mice. (**a**) Cone- and rod-specific ERG responses to light in wildtype (WT, black) and *Gpatch11^Δ^*^5^*^/Δ^*^5^ (red) mice at post-natal days 15, 21, 30, 60, 90, 120, 150, and 180. Bars show mean±SEM from nine mice per genotype. n.s., not significant. * p≤0.05, ** p≤0.01, *** p≤0.001, **** p≤0.0001. (**b**) Spider graph presenting outer nuclear layer (ONL) thickness in wildtype (WT, black) and *Gpatch11^Δ^*^5^*^/Δ^*^5^ (red) mice at 15, 60, 120, and 180 days. Bars represent mean ± SEM from three mice per genotype. * p≤0.05, ** p≤0.01, *** p≤0.001, **** p≤0.0001. (**c**) RNAScope analysis of *Gpatch11* (magenta) mRNA in retina sections. DAPI labels cell nucleus (blue). Scale bars: 50 µm. (**d**) Immunostaining of retina sections with anti-GPATCH11 (magenta) antibody. Nuclei are stained with DAPI (blue). Scale bars: 50 µm. (**e**) Quantification of GPATCH11 protein expression in the ONL, inner nuclear layer (INL), and ganglion cell layer (GCL) in wildtype (WT, black) and mutant *Gpatch11^Δ^*^5^*^/Δ^*^5^ (red) mice. Bars represent mean±SEM from three biological replicates. n.s., not significant. * p≤0.05, ** p≤0.01, *** p≤0.001.

Presence of neurological impairment in the four families of this study encouraged us to investigate the presence of neurological deficits in 1-month-old *Gpatch11^Δ^*^5^*^/Δ^*^5^ mice (the opsin content in photoreceptors and ERG responses consistent with vision) and wild-type littermates. The mice were subjected to novel object recognition (NOR), contextual fear conditioning (CFC), Morris water maze (MWM) tests and the open field test (OFT) to assess episodic memory, associative memory, spatial memory and anxiety-like behaviour, respectively. Both male and female *Gpatch11^Δ^*^5^*^/Δ^*^5^ mice demonstrated an inability to recognize novel objects in NOR experiment (**Fig. 8a**) and significantly shorter context-elicited freezing times during the training and testing phases in CFC experiment (**Fig. 8b**), supporting defective episodic and associative memory, respectively. In contrast, spatial memory and anxiety-like behaviour were not significantly different from those of controls (**Supplementary Fig. 7a, b**).

**Fig. 8:**
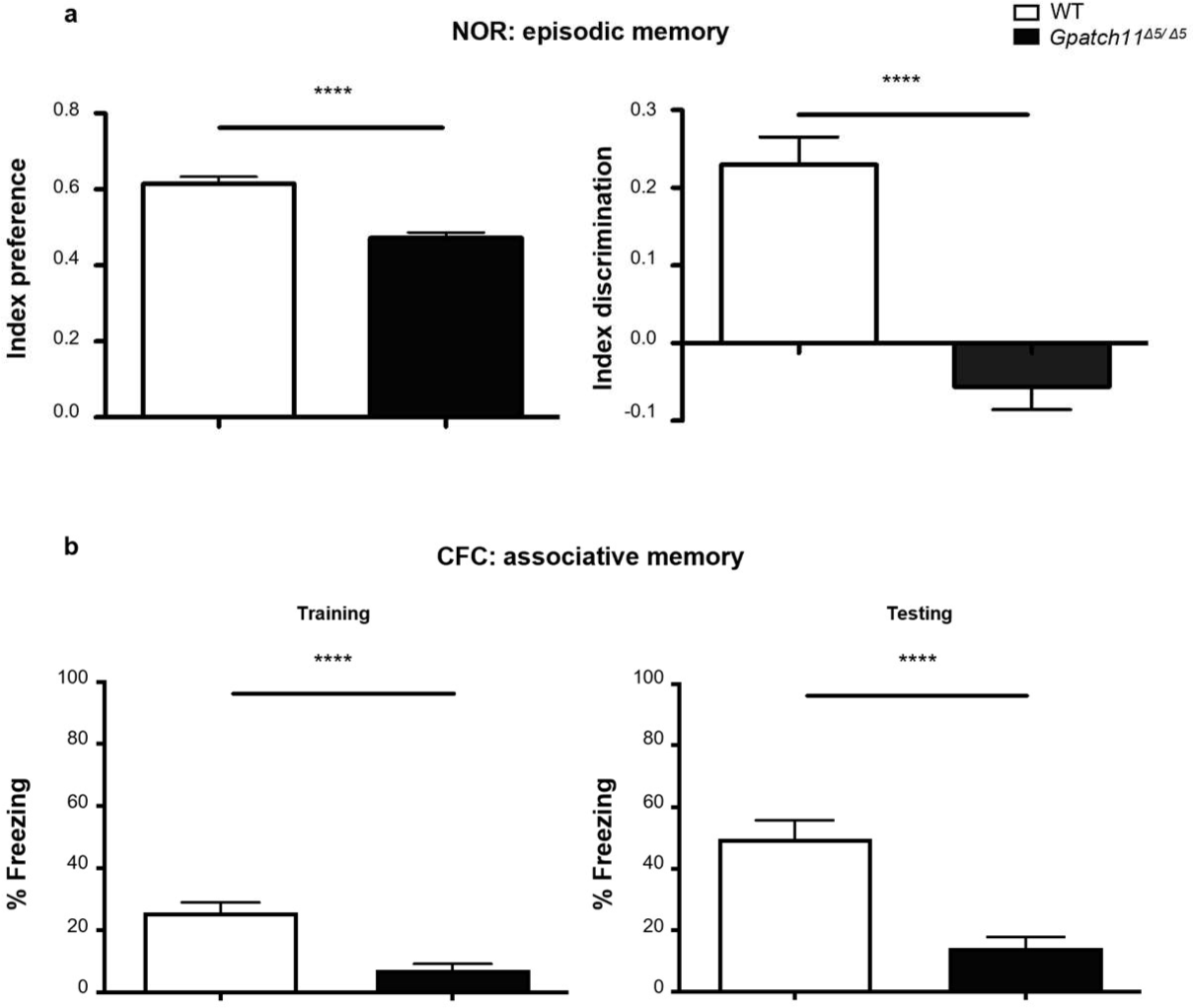
Behavioural tests in 1-month-old wildtype (WT) and *Gpatch11^Δ^*^5^*^/Δ^*^5^ mice. (**a**) Episodic memory assessment based on the Novel Object Recognition (NOR). Preference and Discrimination Indexes measurements. (**b**) Associative memory assessment using the Contextual Fear Conditioning (CFC). Percent Freezing Time index measured during training and testing phases, respectively. Bars represent mean±SEM from 20 mice per genotype. **** p≤0.0001.

Exploration of the retina and the behaviour of our mice model confirmed the presence of the major symptoms described in the human syndrome.

### Wild-type and mutant *Gpatch11* mRNA and protein products are detected in all retinal cells and the mutant protein accumulates in the nucleus of mouse retinal cells

The retinal expression of wild-type and mutant *Gpatch11* mRNA isoforms was examined in eye sections by RNAscope using a probe targeting the two isoforms. Wild-type mRNA in wild-type mice and the mutant counterpart in *Gpatch11^Δ^*^5^*^/Δ^*^5^ littermates were detected and quantified with no significant difference in all retinal layers (**Fig. 7c, Supplementary Fig. 8**).

Immunostaining of the wild-type and mutant protein isoforms in retinal sections using the GPATCH11 antibody showed the presence of the protein in all retinal nuclei (**Fig. 7d**). Of note, we show that in photoreceptor nuclei the protein displayed a perinuclear distribution, contrasting with the nuclear localization observed in other retinal cells. This inverted pattern in photoreceptor nuclei has been previously ascribed to a peculiar nuclear architecture in nocturnal mammalian photoreceptors^16^.

Interestingly, quantification of wild-type and mutant mRNA and protein isoforms over a 6-month period revealed comparable mRNA abundance but significantlly increased GPATCH11 immunostaining in *Gpatch11^Δ^*^5^*^/Δ5^*mice compared to wild-type littermates, suggesting accumulation of the mutant protein (**Fig. 7d**, **e**). Because protein degradation is often modulated by ubiquitination or other post-translational modifications, we investigated posttranslational modifications through proteomic Nextprot database (https://www.nextprot.org/entry/NX_Q8N954/proteomics?isoform=NX_Q8N954-1), which did not identify any modification of the 96-109 amino-acid deleted sequence whose loss could account for altered protein degradation.

Overall, our data show that the human mutant variant in mice produces a shorter GPATCH11 protein, which results accumulated in all the retinal layers while the transcript abundance rests unaltered.

### Expression of the mouse GPATCH11 protein analogous to the human mutant variant affects the splicing and expression of genes related to visual perception in the retina

Whether the loss of the internal portion of the GPATCH-domain of GPATCH11 affected gene expression and/or splicing was investigated by comparing the transcriptomes of retina from 15-day-old *Gpatch11^Δ^*^5^*^/Δ^*^5^ mice and wild-type littermates. This resulted in the identification of 160 differentially expressed genes (DEGs, **Fig. 9a**). GO enrichment analysis indicated the enrichment of genes related to photoreceptors, non-motile cilium and response to light stimuli (phototransduction cascade and visual signal propagation) in the *Gpatch11^Δ^*^5^*^/Δ^*^5^ mice compared to wild-type littermates (**Fig. 9b-d**). Circular visualization based on the calculated z scores indicated that most of the pathways, for which Heatmaps are shown, were enriched in downregulated DEGs in particular (**Fig. 9b, d)**. Neither inflammation nor cell death pathways were dysregulated upon loss of the internal portion of the GPATCH-domain, supporting the view that the transcriptome changes at day 15 were primary consequences of alterations in *Gpatch11*.

**Fig. 9:**
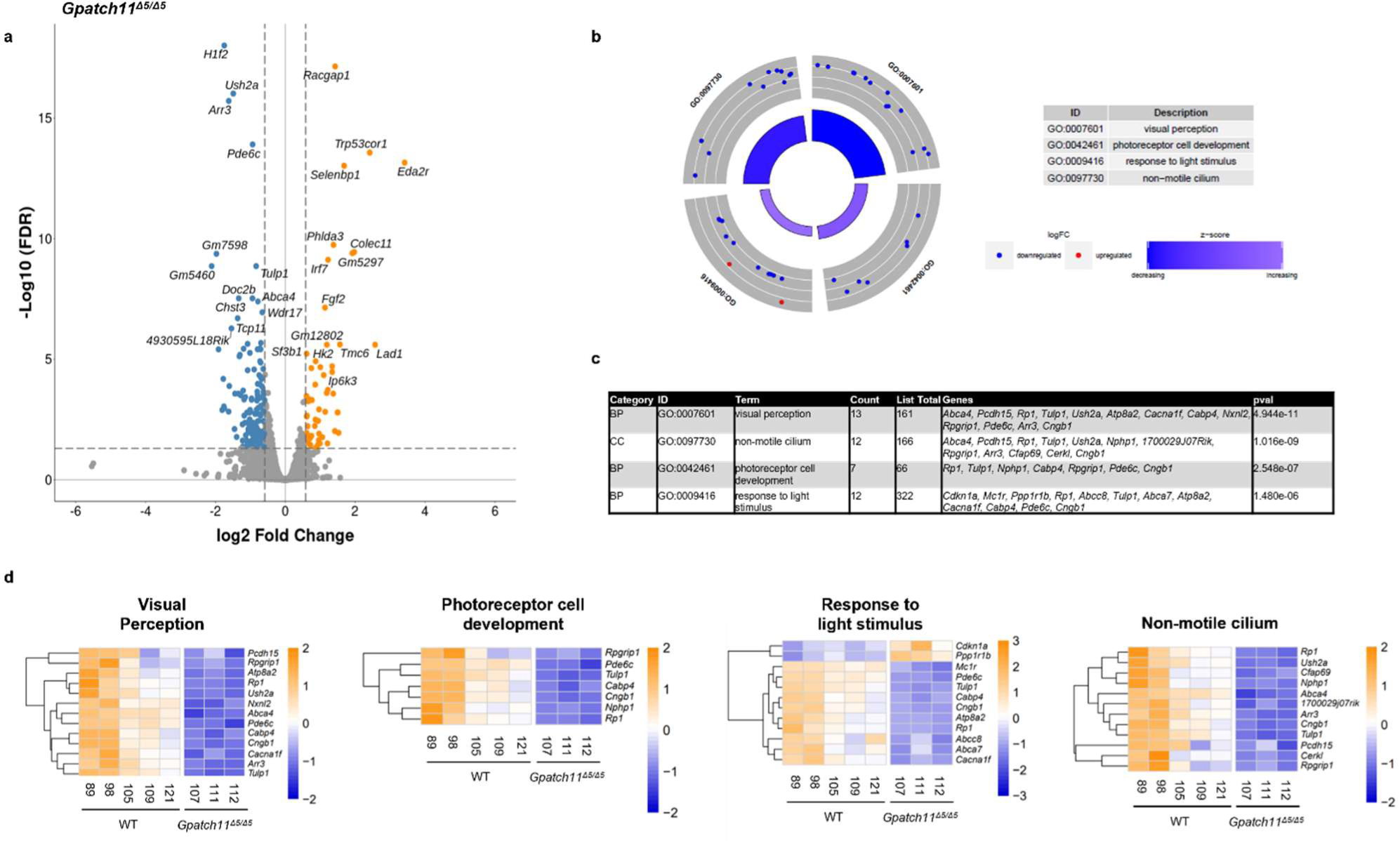
Whole transcriptome analysis of genes expression in retina of wildtype (WT) and *Gpatch11^Δ^*^5^***^/Δ^***^5^ mice. (**a**) Volcano plot displaying differentially expressed genes between *Gpatch11^Δ^*^5^*^/Δ^*^5^ and wildtype (WT) mice. The x-axis represents the difference in gene expression Fold Change (FC) on a log2 scale, while the y-axis indicates False Discovery Rate (FDR) adjusted significance on a log10 scale. Genes significantly upregulated are shown in orange, while downregulated genes are in blue. (**b**) Circular visualization depicting selected Gene Ontology (GO) enriched pathways. Up-(red dots) and down-regulated genes (blue dots) within each GO pathway are plotted based on logFC. Z score bars indicate whether an entire biological process is more likely to be increased or decreased based on its constituent genes. (**c**) Table presenting the over-represented GO pathways of interest identified with Metascape using the Differentially Expressed Genes (DEGs) from the selected clusters. The pathway categories include Biological Process (BP) and Cell Compartment (CC). (**d**) Heatmaps from Metascape displaying the GO pathways of interest. Blue represents low expression, while orange represents high expression. The raw data is accessible via BioStudies identifier S-BSST1157.

Differentially expressed isoform analysis identified 299 splicing events, mainly reduced skipped exons (SEs) and increased mutually exclusive exons (MXEs), in 178 genes (**Fig. 10a**). Differentially spliced genes (DSGs) included genes related to the photoreceptor response to light (phototransduction and synaptic transmission of the visual message), photoreceptor connecting the cilium assembly, protein homeostasis, mitochondria, nucleic acid (chromatin, DNA and RNA) binding and the regulation of RNA splicing (**Fig. 10b**). Most of these genes showed no differences in expression in the DEG analysis, suggesting that splicing events were not responsible for the changes in expression (**Fig. 10c**). Specifically, the intersection of DEGs and DSGs identified only 12 genes: *Arr3, Asap3*, *Cabp4*, *Ccnl2, Dhrs3*, *Lbhd1*, *Mpp4, Pex5l*, *Pitpnm3*, *Tulp1*, *Unc13b*, and *Vtn,* all of which, except *Asap3*, were downregulated upon loss of the internal portion of the GPATCH-domain (**Fig. 10c, d**). The splicing events in *Asap3, Cabp4, Lbhd1, Pitpnm3, Tulp1,* and *Vtn* are expected to introduce premature termination codons in the mRNA and trigger nonsense-mediated mRNA decay. In contrast, analysis of the splicing events in *Arr3*, which is reported as an example, suggests that mis-splicing is not the main driver of expression dysregulation in this gene (**Fig. 10e-g**) as well as in *Ccnl2, Dhrs3, Mpp4, Pex5l,* and *Unc13b* (**Supplementary Table 3**).

**Fig. 10:**
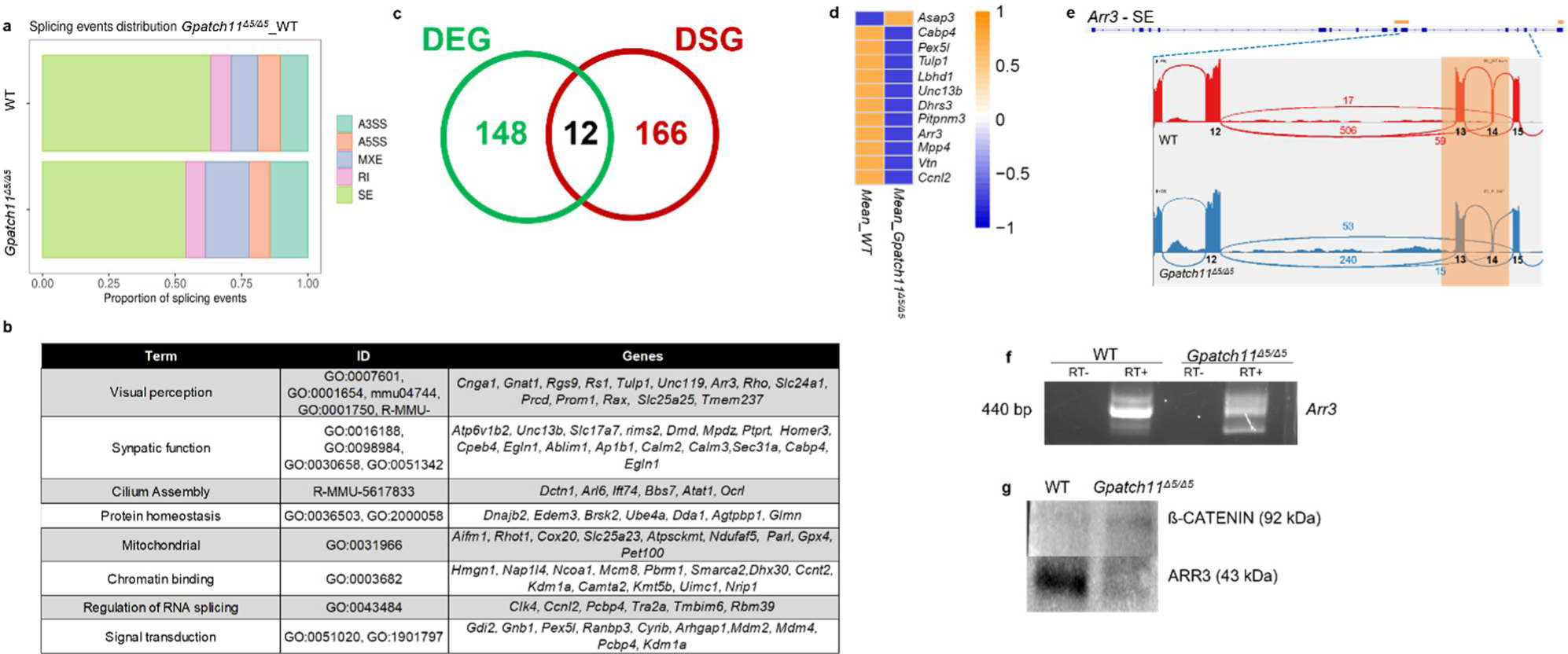
Whole-transcriptome analysis of splicing in the retina of wildtype (WT) and *Gpatch11^Δ^*^5^*^/Δ^*^5^ mice. (**a**) Chart generated from rMATS analysis, illustrating that the majority of splicing events corresponded to skipped exons. A5SS and A3SS represent alternative 5′ and 3′ splice sites, SE denotes skipped exons, RI stands for retained introns, and MXE indicates mutually exclusive exons. (**b**) Table displaying the over-represented Gene Ontology (GO) pathways of interest, identified using the Differentially Spliced Genes (DSGs) from the selected clusters with Metascape. (**c**) Venn diagrams comparing Differentially Expressed Genes (DEG) and DSG in *Gpatch11^Δ^*^5^*^/Δ^*^5^ as compared to wildtype (WT) retina samples, with a total of 12 dysregulated and mis-spliced transcripts. (**d**) Heatmaps depicting the expression levels of the 12 dysregulated and mis-spliced transcripts. Blue indicates low expression, while orange represents high expression. (**e**) Sashimi plots illustrating the alternate splicing event for *Arr3* in retina samples from wildtype (WT, blue) and *Gpatch11^Δ^*^5^*^/Δ^*^5^ (red). Orange highlights in the sashimi plots indicate the alternative splicing events, and numbers indicate the number of junction reads for each event. The raw data can be accessed via BioStudies and the identifier S-BSST1157. (**f**) Electrophoresis of *Arr3* cDNA and (**g**) Western blot analysis and ARR3 protein relative to β-CATENIN in wildtype (WT) and *Gpatch11^Δ^*^5^*^/Δ^*^5^ retina samples.

### Expression of the mouse GPATCH11 protein analogous to the human mutant variant affects the expression of proteins related to visual perception and to spliceosome in the retina

Mass spectrometry (MS) analysis of total-retina lysates from 21-day-old *Gpatch11^Δ^*^5^*^/Δ^*^5^ mice and wild-type littermates detected 150 proteins in the *Gpatch11^Δ^*^5^*^/Δ^*^5^ mice with either decreased (n=63/150) or increased (n=87/150) abundance. The downregulated proteins were involved in several of the pathways identified by transcriptome analysis, namely, the response to light stimulation, synaptic transmission of the visual message, mitochondria, cilia, protein homeostasis, and RNA binding and splicing (**Fig. 11a**). Nine of these proteins were encoded by downregulated DEGs, namely, *Arr3, Cabp4*, *Eml3, Lhbd1, Nxnl2, Mpp4, Pex5l*, *Smad11*, and *Tulp1* (**Fig. 11b**). Among them, *Tulp1* and *Cabp4* have been found to be associated with LCA15^17^ and cone-rod synaptic disorder (CRSD)^18^, respectively, and *Arr3* encodes cone arrestin, a key player in the light-dark adaptation of cones^19^. Among the 54 remaining proteins in this group, 12/54 are involved in RNA processing, and 8/12 are involved in splicing and present in spliceosomal complexes: SNRPA (in U1), DDX17 and FUS (in Complex A), DDX42 and DNAJC8 (in U2), FAM32A (in Complex C), HNRNPM and HNRNPH3 (in hnRNP) (**Fig. 11a**). Among the 87 proteins upregulated in *Gpatch11^Δ^*^5^*^/Δ^*^5^ mice, 3 were encoded by genes with increased mRNA abundance (e.g., *Fgf2, Apobec2, Txnl4a;* **Fig. 11c, d**). The remaining 84 proteins are encoded by genes whose RNA expression was similar in *Gpatch11^Δ^*^5^*^/Δ^*^5^ mice and wild-type littermates. GPATCH11 was included in this list of these 84 upregulated proteins, consistent with immunostaining studies showing accumulation of the mutant product in the retinas of *Gpatch11^Δ^*^5^*^/Δ^*^5^ mice (**Fig. 11e**).

**Fig. 11:**
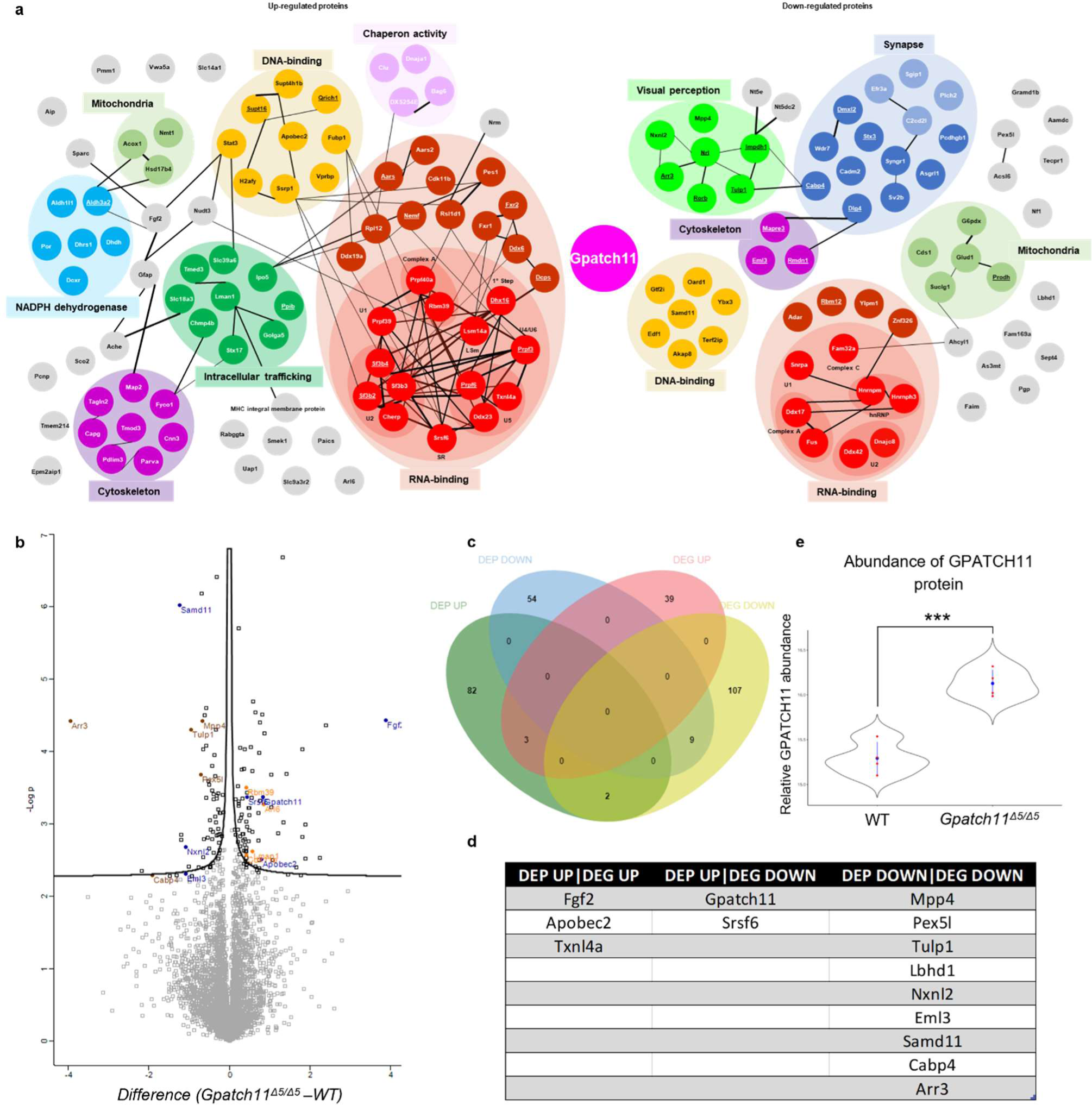
Mass spectrometry analysis of retina lysates from wildtype (WT) and *Gpatch11^Δ^*^5^*^/Δ^*^5^ mice. (**a**) Schematic representation depicting protein interactions found in the total lysate of *Gpatch11^Δ^*^5^*^/Δ^*^5^ mouse retinas. (**b**) Volcano plots illustrating differentially expressed proteins in *Gpatch11^Δ^*^5^*^/Δ^*^5^ mice as compared to wildtype (WT) counterparts. (**c**) Venn diagrams comparing up-regulated and down-regulated differentially expressed genes (DEG) and differentially expressed proteins (DEP) in *Gpatch11^Δ^*^5^*^/Δ^*^5^. (**d**) Table of genes identified in the intersection of the Venn diagram. (**e**) Violin plot representing the abundance of the GPATCH11 protein. The raw data is accessible via ProteomeXchange with the identifier PXD041849.

GO enrichment analysis showed that RNA-binding proteins represented more than half of the overrepresented proteins, including PRPF39 (in U1), CHERP, a G-patch-domain-containing protein that is known to play a role in pre-mRNA splicing; SF3B2; SF3B3 and SF3B4 (in U2); PRPF3 (in U4/U6); DDX23; PRPF6 and TXNL4 (in U5); PRPF40A and RBM39 (in Complex A); DHX16 (in 1^st^ Step); SRFS6 (in SR); and LSM14A (in LSm) (**Fig. 11a**). Several down- and upregulated spliceosomal proteins were reported to be involved in nonretinal (TXNL4^20^, SF3B4^21^ and FUS^22^) and retinal-specific (PRPF3^23^ and PRPF6^24^) spliceosomopathies, respectively^1^.

Together, these results are consistent with a role for GPATCH11 in splicing and the transcriptional regulation of genes important to retinal function, and possibly other functions through the dysregulation of nonretinal spliceosomopathy-associated genes.

## DISCUSSION

In recent years, G-patch-domain-containing proteins have emerged as pivotal regulators of RNA metabolism, assuming diverse roles that encompass pre-mRNA splicing and transcriptional regulation. The bulk of our knowledge about GPATCH protein functions has been derived from *in-vitro* studies conducted in yeast and human cell models^2,3^. Surprisingly, the study of rare genetic diseases has made only minimal contributions to our understanding of GPATCH functions. To date, *RBM10*^7^ and *SON*^8^ are the two GPATCH genes with demonstrated mutations causing human disease.

In this report, we present compelling evidence supporting the notion that biallelic mutations in *GPATCH11*, one of the lesser-known members of this family^2^, underlie a syndromic disease in four families. The families initially sought genetic consultation due to distinct and varying presenting symptoms, reflecting the variable severity of retinal disease. Notably, retinal impairment, was the primary symptom in Family 1, while intellectual disability took precedence in the other pedigrees. Moreover, various clinical features, including seizure, short stature, and diabetes, were associated with this retinal disease. It’s worth mentioning that these symptoms were reported in the eldest individuals available for examination in the series, and evaluation in the youngest affected subjects is pending.

Additional symptoms linked with *GPATCH11* mutations are possible. This is highlighted by the sudden death during teenage years and young adulthood of two unrelated individuals from a fever episode, which could indicate defective responses to pathogens, and thalamic stroke, suggesting a need for specific investigations into immune responses among individuals carrying *GPATCH11* disease-causing variations.

Considering the severe phenotype affecting male gonads in the mice, it would be worthwhile to examine the fertility of affected male individuals in Families 1, 2 and 4.

Notably, genotype–phenotype correlation analysis revealed no significant difference in disease presentation between individuals who were homozygous for the splice site variant (Families 1 and 3), subjects carrying a nonsense mutation *in trans* with it (Family 2) or the individual carrying a frameshifting splice site mutation homozygously (Family 4). Although the frameshift is predicted to lead to absence of the protein, the possibility of partial bypassing of protein truncation through self-correcting mechanisms cannot be excluded. To validate the effect of the predicted frameshift splice site mutation, analysis of the RNA derived from the cells of patient from Family 4 would be valuable.

While investigating the subcellular localization of GPATCH11 in human fibroblasts, we made intriguing observations about the nuclear distribution of the protein which displayed a distribution pattern characterized by nuclear speckles and a diffuse presence in the nucleoplasm, a pattern reported for several spliceosome components^10,11^. Consistent with a role for GPATCH11 in the spliceosome, we observed a significant reduced abundance of U4 snRNA, and possibly U5 snRNA, in fibroblasts homozygous for a *GPATCH11* mutation identified in 3 out of 4 families. This mutation resulted in a protein isoform lacking the central part of the GPATCH-domain while keeping the other domains intact. Notably, the U4 and U5 snRNAs participate with the U6 snRNA to a complex known as the U4/U6.U5 tri-snRNP^25^. It is interesting to note that mutations in the U4/U6-specific protein PRPF31^26^, the U5 protein PRPF6^24^ as well as PRPF3^23^ and PRPF4^27^ which are all components of the U4/U6.U5 tri-snRNP complex have been linked to retinal dystrophy^1^.

Additionally, our analysis of the retina of mice expressing a GPATCH11 protein analogous to the human mutant variant, which faithfully recapitulated the retinal disease and neurological abnormalities associated with human *GPATCH11* mutations, unveiled highly significant splicing abnormalities. Interestingly, we also detected dysregulation of gene expression, which, like splicing defects, impacted mainly photoreceptor light responses which dysregulation is known to trigger photoreceptor cell death, leading to visual loss^28^. Reduced mRNA abundance could be a direct consequence of splicing aberrations leading to the degradation of mRNA isoforms containing premature termination codons or an indirect result of splicing dysregulation. However, we identified certain genes, such as *Arr3*, which exhibited reduced mRNA levels and abnormal splicing without altering the reading frame. This finding suggests that, akin to other G-patch-domain-containing proteins like SON and ZGPAT^2^, GPATCH11 is involved in both pre-mRNA splicing and transcriptional regulation.

In mutant mouse retina, we observed GPATCH11 accumulation, which might result from the loss of residues essential for its degradation or be an indirect consequence of a broader dysregulation^29^. This accumulation led to the massive dysregulation of RNA-binding protein complexes, most of which also showed increased accumulation. Several are known to be involved in the spliceosome and mutations in their components have been linked to non-retinal (TXNL4A^20^) and retinal-specific (PRPF3^23^, PRPF6^24^) spliceosomopathies.

Very interestingly, alongside with a nuclear distribution, we observed GPATCH11 in the centrosome. The nuclear and centrosomal/cilia distribution of GPATCH11 aligns with recent findings describing it as a characteristic feature of spliceosome proteins known as centrosome-associated spliceosome components emerging as crucial players in ciliogenesis and tissue specification, reshaping our understanding of RNA metabolism within the cell^11^. Until recent years, the conventional belief that pre-mRNA splicing exclusively takes place within the nucleus faced challenges, primarily due to reports of centrosome/cilia enrichment of both non-coding and coding RNA, along with dedicated translation machinery. Furthermore, the localization of spliceosomal components to these cellular regions further questioned this traditional view. This centrosome-associated RNA machinery is hypothesized to facilitate the nucleocytoplasmic shuttling of RNA between the spliceosome and the cytoplasm^11^.

The precise function of RNA-processing complexes at the centrosome remains an ongoing area of investigation. One possibility is that cell type-specific splicing factors assemble on RNA and congregate around the ciliary basal body, thereby enhancing ciliogenesis and the cellular response to stimuli. A similar concept can be seen in the local translation of nuclearly encoded mitochondrial mRNAs, which facilitates the rapid on-site supply of mitochondrial membrane proteins^30^. The existence of a close relationship between the spliceosome and the centrosome is evident in the striking phenotypic similarities between splicing and centrosome-related genetic disorders, particularly retinal degeneration, brain, and craniofacial developmental defects observed in individuals carrying *GPATCH11* mutations.

The question of whether GPATCH11’s presence at the cilia basal body contributes to cilia-related functions remains unanswered. Our investigation into cilia formation and SHH signalling in fibroblasts from patients did not uncover any abnormalities. However, it’s essential to consider that this lack of findings may be attributed to the specific cell type under examination.

Adding support to this perspective is the noteworthy dysregulation in the expression of primary cilium proteins in the retina of mutant mice. These proteins are integral components of the photoreceptor connecting cilium and outer segments, and their dysregulation have been largely reported to contribute to the observed retinal phenotype. Among them *Tulp1* has been found to be associated with LCA15^17^. This suggests that while our fibroblasts analysis may not have yielded apparent abnormalities, the impact of GPATCH11 mutations on ciliary functions may manifest differently in other cell types or tissues, emphasizing the need for further investigations to comprehensively unravel the intricacies of GPATCH11’s role in cilia-related processes.

Interestingly, the retina of the mutant mice developed and layered normally, responding appropriately to light at eye opening, and degenerated posteriorly with light exposure. These findings suggest that GPATCH11 may play a crucial role in the function and maintenance of photoreceptor cells but not in the initial development of the retina. The variability in the onset and severity of retinal disease in affected individuals from our cohort further supports this hypothesis.

The origin of the neurological anomalies remains ambiguous. It is unclear whether they result from developmental issues or arise later in life, particularly considering that neurological problems in the affected patient of Family 1 manifested rather late in childhood. Conversely, the presence of dysmorphic features in patients provides evidence that GPATCH11 serves essential functions both during early tissue development and in the subsequent maintenance of proper function.

While both ciliopathies and spliceosomopathies exhibit features involving the retina, brain, and skeletal system, some distinct differences have emerged, particularly in the context of male sterility in mouse models. To our knowledge, thus far male sterility has been associated with ciliopathies but not spliceosomopathies.

Furthermore, there are additional clinical manifestations in patients such as seizures, diabetes, and possibly immune deficiency that are typically not observed in either of these disease families.

This raises intriguing questions about the broader functions of GPATCH11 in RNA homeostasis, extending beyond a role in pre-mRNA splicing and ciliary function. Could GPATCH11’s involvement in these broader RNA-related processes explain its association with a more diverse spectrum of clinical phenotypes compared to traditional spliceosomopathies and ciliopathies?

To gain deeper insights into these questions, further research is clearly warranted. Investigating GPATCH11 intricate roles in ciliary function and its implications for the wide range of clinical manifestations associated with *GPATCH11* mutations is essential.

In summary, our research uncovers a complex syndrome resulting from *GPATCH11* mutations, characterized by clinical features overlapping spliceosomopathies and ciliopathies.

Furthermore, our investigation indicates that GPATCH11 plays a pivotal role in pre-mRNA splicing and in gene transcription regulation, particularly for genes crucial in the function and maintenance of rod and cone photoreceptor cells, as well as brain development. These collective findings shed light on the intricate and diverse roles of GPATCH11 in RNA metabolism, potentially extending to cilia metabolism, underscoring its significance in various physiological processes. They provide valuable insights into the molecular underpinnings of the clinical features associated with *GPATCH11* mutations. Moreover, our research offers a model that could contribute to our understanding of how RNA processing contributes to ciliogenesis and genetic disorders, marking an enticing avenue for future research in this field.

## MATERIAL AND METHODS

### Families

The study involved ten affected subjects (six females and four males) with retinal dystrophy from four families, three of which are multiplex and consanguineous (Family 1 and 3), one is a multiplex and non-consanguineous (Family 2) and one is non-multiplex and consanguineous (Family 4). Family 1 was referred for early-onset retinal dystrophy with neurodevelopmental delay and consists in an inbred north African pedigree with several loops of consanguinity and four affected individuals (two affected children and two affected relatives, one of whom deceased). Family 2 was referred for intellectual disability and comprises three affected individuals, one sibling and their two unrelated parents from North Africa. Family 3 includes two affected subjects born to consanguineous parents from North Africa and had been reported previously^31^. Family 4 includes one affected individual from Europe. All individuals or legal representatives consented with the study, which received approval from the institutional review boards Comité de Protection des Personnes Ile de France II (Necker), Reference Center for Congenital Abnormalities and Malformative Syndromes in Dijon (France) and *Orphonomix units* for genetic testing, located in several hospitals in France. All participating individuals of Family 4 signed a consent form, in agreement with the Declaration of Helsinki and the ARVO statement on human subjects. Genomic DNA was extracted from peripheral blood by standard procedures.

### Gene-panel testing and exome sequencing (ES)

The *GPATCH11* variants were identified by IRD panel followed by whole-exome sequencing (WES) in research settings (Family 1) or in diagnostic settings in clinical laboratories and direct WES (Families 2, 3 and 4). Genomic DNA libraries were generated from DNA (F1:III-6; F1:IV-1; F1:IV-2; F2:I-1, F2:I-2; F2:II-2) sheared with a Covaris S2 Ultrasonicator via SureSelect^XT^ Library Prep Kit (Agilent). Regions of interest (ROIs) were captured with the SureSelect All Exon V5 kit (Agilent) and sequenced on an Illumina HiSeq2500 HT system (Illumina). Data analysis was performed with a homemade pipeline (POLYWEB)13 created by the *Imagine* Institute Bioinformatics core facilities of Paris Descartes University. The following algorithms were used to predict the consequences of variants identified with WES: PolyPhen-2, SIFT and MutationTaster. Allele frequencies were evaluated via the gnomAD population database. The Segregation analysis was performed in parents of affected individuals via Sanger sequencing. It demonstrated biparental transmission in all the affected individuals. WES and variant filtration in Family 3 has been previously described^31^. WES analysis of individuals from Family 4 was performed at Otogenetics (Norcross, GA) using Agilent V4 (Santa Clara, CA, USA) and Illumina HiSeq 2000 with 30 × coverage. Variants in *GPATCH11* were annotated using GRCh38 reference genome and GenBanck transcript (NM_174931.4).

### GPATCH11 variant identification

GPATCH11 variants were identified through a two-step process: initially via an IRD panel followed by whole-exome sequencing (WES) in research settings (Families 1 and 4) and in diagnostic settings at clinical laboratories through direct WES (Families 2 and 3).

The whole exome of F1:IV-1 (P1), F1:IV-2 (P2), F1:III-6 (P4), F2:I-1, F2:I-2, F2:II-2 (P5) individuals from Families 1 and 2, respectively, was sequenced as described32. In Brief, DNA libraries were prepared using the SureSelect^XT^ Library Prep Kit (Agilent Technologies). Regions of interest were captured using the SureSelect All Exon V5 kit (Agilent Technologies) and run in pair-ended mode (2 × 75) on an Illumina HiSeq2500 HT system (Illumina). Sequences were aligned to the human genome reference sequence (UCSC Genome Browser hg19 assembly). Genetic variation annotation and variant filtration were performed using the POLYWEB (https://polyweb.fr/) pipeline developed by the *Imagine* Institute Bioinformatics core facility and Paris Descartes University. Within the variant filtration setting of POLYWEB, various parameters are integrated. This includes variant frequencies based on gnomAD data, pathogenicity scores derived from reference predictors (including PolyPhen, SIFT, CADD, splice AI), and the ability to attribute genetic models. WES and variant filtration in Family 3 have been previously documented^31^. For Family 4, WES was conducted at CeGaT GmbH in Tübingen, Germany. Here, sequencing libraries were generated using the Twist Human Core Exome Plus kit (Twist Bioscience) following the manufacturer’s protocols. Paired-end sequencing on a Novaseq 6000 produced 100-base sequences. Variant filtration and prioritization were performed using an in-house pipeline^33^. Variants in *GPATCH11* were identified by selecting rare sequence changes (minor allelic frequency >1%) with highest scores of pathogenicity and consistency with a recessive model of inheritance. Variants have been annotated using the GRCh38 reference genome and GenBank transcript (NM_174931.4).

### Analysis of c.328+1G>T and c.454C>T variants on mRNA

A skin biopsy was obtained from the F1:IV-1 (P1) and F1:IV-2 (P2) probands and four controls (C1-4). Primary fibroblasts were isolated by selective trypsinization and proliferated at 37°C, 5% CO2 in Opti-MEM GlutaMAX I medium supplemented with 10% FBS (Fetal Bovine Serum), 1% streptomycin/penicillin (Thermo Fisher Scientific). Genomic DNA and total RNA were extracted from trypsinized cells using the Quickextract DNA and RNAeasy Mini kits, according to the manufacturers’ protocols (Lucigen and Qiagen, respectively). RNA from peripheral blood white cells of patient (F2:II2, P5) carrying the c.328+1G>T and c.454C>T variants in compound heterozygosity was prepared using the PAXgene Blood RNA Kit (50) v2 protocol (Qiagen). All samples were DNase treated by the RNase-free DNase set (Qiagen). Concentration and purity of total RNA was assessed using the Nanodrop-8000 spectrophotometer (Thermo Fisher Scientific). First-stranded cDNA synthesis was performed from total RNA (500 ng) using Verso cDNA kit (Thermo Fisher Scientific) according to the manufacturer’s instructions. A non-reverse transcriptase (RT) reaction (without enzyme) for one sample was prepared to serve as control in RT-PCR experiment.

For analysis, gDNA (50 ng) and cDNA (5 µL of a 1:25 dilution in nuclease-free water) were PCR amplified in a 5Xbuffer (2μl) containing 1 µM of specific primers to *GPATCH11* (**Supplementary Table 2**), dNTP (10 µM), MgCl2 (2.5 mM) and DNA polymerase (0.5 units GoTaq, Promega).

PCR products (2 μl) were subjected to a sequencing reaction using BigDye Terminator v3.1 Cycle Sequencing Kit, according to the recommendation of manufacturer (Applied Biosystem). Purified products were sequenced on a 350xL Genetic Analyzer ABI and analyzed using the Sequencing Analysis v5.2 sequence software.

### Quantitative analysis of U1, U2, U4, U5, U6 snRNA abundance in patient and control fibroblasts

snRNAs were extracted from F1:IV-1 (P1), F1:IV-2 (P2) and control fibroblasts using the Trizol and digested through DNAse RQ1 treatment, according to the manufacturer’s protocols (Qiagen and Promega, respectively). Assessment total RNA concentration and purity and first-stranded cDNA synthesis were performed as described previously. A non-reverse transcriptase (RT) reaction (without enzyme) for one sample was prepared to serve as control in RT-qPCR experiment.

For RT-qPCR analysis, cDNAs were amplified using specific primers designed from the sequences of U1, U2, U4, U5, U6 snRNA and *HPRT1* and *RPLP0* housekeeping genes. All primers have been previously reported^34,35^ but the U6 snRNA specific primers: Forward 5’-GCTTCGGCAGCACATATACTAAAAT-3’, reverse 5’-CGCTTCACGAATTTGCGTGTCAT-3’. cDNAs (5 µL) were subjected to real-time PCR amplification in a buffer (20 µL) containing SYBR® Green Master mix (Applied Biosystems) and 0,5 μM of forward and reverse primers, on a mastercycler realplex2 (Eppendorf). Data were analysed using the realplex software (Eppendorf). For each cDNA sample, the mean of quantification cycle (Cq) values was calculated from triplicates (SD<0.5 Cq). Expression levels U1, U2, U4, U5, U6 snRNAs were normalized by using the relation ΔΔCq relative to HPRT1 and RPLP0. Absence of amplification when using mRNA (non-RT) and water (W) as templates were controlled in each run (Cq values non-RT >30 and W=undetermined). The quantitative data are the means ± SEM of three independent experiments and these are presented as ratio among values for individual mRNAs. GraphPad Prism 6 software was used for statistical analyses. The significance of variation among samples was determined using Student’s t test. Error bars reflect the standard error of the mean (SEM).

### Western blot analysis of GPATCH11

Cells were incubated in the radioimmunoprecipitation assay (RIPA) lysis buffer (Thermo Fisher Scientific) for 1 hour in ice, sonicated for 20 seconds (1 pulse at 70% of amplitude) and quantified using the Bradford assay. Proteins (50 µg) were denatured at 95°C for 8 minutes in a solution (20 μl) containing LDS Sample Buffer 1X (Invitrogen) and 10% of 2-Mercaptoethanol and run in a Mini-PROTEAN TGX Stain-Free Any kD gel, according to the recommendations of the supplier (BioRad). Separated proteins were transferred onto a PVDF membrane using the TransBlot Turbo Mini-size PVDF membrane system, following the manufacturer recommendations (BioRad). The membrane was incubated overnight at 4°C with the polyclonal rabbit anti-CCDC75 (anti-GPATCH11) primary antibody raised against the antigenic sequence encompassing amino acids 111-192 of the human protein (Abcam) (1/4000). The goat anti-rabbit IgG-HRP was used as secondary antibody (Invitrogen; 1/4000). Immunoblots were revealed using Clarity Western ECL Substrate (BioRad) and the ChemiDoc XRS+ Imagin System (BioRad). Western blot images were acquired and analyzed with Image Lab software 3.0.1 build 18 (Bio-Rad). To compare GPATCH11 abundance across samples, membranes were secondarily incubated with mouse anti-β-ACTIN and goat anti-mouse IgG-HRP primary and secondary antibodies, respectively. The abundance of GPATCH11 relative to ß-ACTIN was estimated in each cell line by densitometry using Image Lab software (BioRad). The quantitative data are the means±SEM of three independent protein extractions and these are presented as ratio relative to β-ACTIN. Data were analysed using GraphPad Prism 6 software. The significance of variations among samples was estimated using the one-way ANOVA with a post hoc Tukey’s test.

### Immunocytochemistry analysis of *GPATCH11* subcellular localization

Primary cultured fibroblasts from F1:IV-1 (P1), F1:IV-2 (P2) and control fibroblasts were seeded at 2×10^5^ cells/well on glass coverslip in 12-well or in 6-well plates and maintained in at 37°C, 5% CO2 in Opti-MEM GlutaMAX I medium supplemented with 10% FBS (Fetal Bovine Serum), 1% streptomycin/penicillin (Thermo Fisher Scientific) for 24h. Cells were then fixed with PFA (4%) or methanol (100%) and blocked with bovine serum albumin (BSA) 3% and triton 0.1% in phosphate buffered saline (PBS). GPATCH11 was immunostained overnight at 4°C using the polyclonal rabbit anti-GPATCH11 (1:400; Abcam) and co-immunostained with the monoclonal mouse anti-SC-35 (1:2000, Sigma-Aldrich), the monoclonal mouse H3K9me3 (1:500, Active Motif) and the monoclonal mouse γH2AX (1:500, Millipore) antibodies. The primary antibodies were labelled for 1 h at room temperature using the Alexa 555-conjugated donkey anti-rabbit (1:1000; Life Technologies) and the Alexa 488-conjugated donkey anti-mouse (1:1000; Life Technologies) secondary antibody. Nuclei were stained using DAPI (Invitrogen). Images were recorded from Spinning Disk Zeiss microscope (Zeiss) using a 40x/1.3 Oil objective. For the analysis of GPATCH11 immunostaining, we first used a machine learning with Ilastik^12^ (v1.3.3post3) for GPATCH11 and Cellpose2^36^ nucleus detection. Then, means intensities of nuclear and cytoplasmic GPATCH11 staining we measured with Fiji^37^ macro. Image figures were made through FigureJ^38^. GraphPad Prism 6 software was used for statistical analyses. The mean intensity of nuclear and cytoplasmic GPATCH11 in fibroblasts of affected individuals (Ps) and controls (Cs) was compared using a one.way ANOVA with a post-hoc Tukey’s test.

Due to an unforeseen centrosomal detection of GPATCH11, the subcellular localization of the protein was further analyzed by immunocytochemistry of serum-starved cells to promote ciliation. The cell were fixed in methanol (100%) and immunostaining using the polyclonal rabbit anti-GPATCH11 (1:400; Abcam) antibody and the mouse monoclonal anti-α-TUBULIN ACETYLATED (1:1000, Sigma-Aldrich; axonemal staining), anti-ROOTLETIN (1:200, Santa Cruz Biotechnology; centrosome linker), anti-CENTRIN3 (1:200, Abnova; mother centriole), primary antibodies and ATTO 550-conjugated goat anti-rabbit (1:700; Sigma-Aldrich) and Alexa 514-conjugated goat anti-mouse (1:700; Life Technologies) secondary antibodies, respectively. Nuclei were stained using DAPI (Invitrogen). The images were scanned with the Confocal Leica SP8 gSTED (Leica) and final images were analyzed with Fiji^37^.

### Analysis of cilia formation in fibroblast cells

Primary cultured fibroblasts from F1:IV-1 (P1), F1:IV-2 (P2) and control fibroblasts were fixed with methanol 100% and blocked with bovine serum albumin (BSA) 3% and triton 0.1% in phosphate buffered saline (PBS). Ciliary axoneme and basal bodies were stained using mouse monoclonal anti-acetylated α-TUBULIN (Sigma Aldrich; 1:2000) and rabbit polyclonal anti-PERICENTRIN (1:5000, Abcam) primary antibodies overnight at 4°C, and Alexa 488-conjugated donkey anti-mouse (1:1000 Life Technologies) and Alexa 555-conjugated donkey anti-rabbit (1:1000; Life Technologies) secondary antibodies for 1 h at room temperature, respectively. Images were recorded from Spinning Disk Zeiss microscope (Zeiss) using a 40x/1.3 Oil objective. Mean numbers of ciliated cells were calculated from >100 cells each from the four individual controls, in three independent experiments. Cilia lengths were measured from the same immunofluorescent images. Mean values were calculated from >100 cells in three independent experiments for each cell line for each cell line. Data were analysed using GraphPad Prism 6 software. Data from patient and control cells were compared using the Student’s t test. Error bars reflect the standard error of the mean (SEM).

### RT-qPCR analysis of SHH signaling in fibroblast cells

Patient and control fibroblasts were subjected to a 48-hour period of serum starvation. Subsequently, they were either exposed to a smoothened agonist (SAG, 100 nM) or left untreated as a negative control for a duration of 24 hours. RNA was separately extracted for each condition and then converted into cDNA, following previously described procedures. The cDNAs were then amplified using specific primers designed based on the sequences of *GLI1*, *GLI2*, *SMO*, and *PTCH1*, along with the housekeeping genes *GUSB* and *HPRT1*^35^.Quality control measures were implemented, including verifying the absence of amplification when using mRNA (non-RT) and water (W) as templates in each run. RTq-PCR conditions, data analysis, and statistical analysis of variations among the samples were performed using one-way ANOVA through post hoc Sidak’s test. GraphPad Prism 6 software was used for statistical analyses. Error bars reflect the standard error of the mean (SEM).

### Generation and analysis of genome-edited hTERT-RPE1

To establish a cell line expressing a GPATCH11 protein with its CCDC-domain removed while keeping the other domains intact, we employed the CRISPR-Cas9 genome editing strategy, as previously outlined in reference^39^. In brief, we designed two guide RNAs (sgRNAs) with specific targeting for the in-frame GPATCH11 exon 3, which encodes the CCDC-domain. These sgRNAs, namely Guide 5’1 (5’-TGCCTTAGCATTGGCAATCCTGG-3’) and Guide 3’1 (5’-GAACAAGAAAGACGTGACATTGG-3’), were designed for exon 3 using the CRISPOR software. The selection was based on their on-target and off-target scores, accessible at (http://crispor.tefor.net/).

The CRISPR-Cas9/sgRNA ribonucleoprotein (RNP) complex was created by annealing a crRNA XT recognition domain with an ATTO 550-tagged tracrRNA transactivator domain (Integrated DNA technologies) and assembling with the S. pyogenes HiFi Cas9 Nuclease V3 nuclease (Integrated DNA technologies), following the manufacturer protocol. These RNP complexes were mixed with hTERT-RPE1 cells and introduced into a 16-well reaction cuvette using the 4D-Nucleofector System (Lonza). The cells were nucleofected utilizing program CA137 on the 4D-Nucleofector system. After 24 hours, cells displaying ATTO 550 fluorescence were sorted via flow cytometry (BD FACS ARIA II SORP, BD Biosciences) and subsequently placed into 96-well plates for single-cell selection. Cell clones were propagated on 96-well plates in Dulbecco modified eagle’s medium (DMEM F-12), supplemented with 10% FBS (Fetal Bovine Serum) and 1% streptomycin/penicillin (Thermo Fisher Scientific). Upon cell passage, a portion of the cells was used for DNA extraction and PCR amplification, employing intronic primers flanking exon 3 (**Supplementary Table 2**). The PCR products were separated by agarose gel (2%) electrophoresis to identify samples displaying a unique band, the molecular weight of which was consistent with the homozygous deletion of exon 3. PCR products from cells meeting this criterion were subsequently subjected to Sanger sequencing, as described previously. Total RNA was extracted from trypsinated cells using the RNAeasy Mini kits, according to the manufacturer’s protocols (Qiagen). Assessment total RNA concentration and purity and first-stranded synthesis were performed as described previously. Cells were incubated and lysed with RIPA-PIC (Thermo Fisher Scientific) and proceeded further for Western blot analysis as described previously.

### Immunocytochemistry analysis of GPATCH11 subcellular localisation and cilia formation analysis in genome-edited hTERT-RPE1

To analyse the subcellular localisation of GPATCH11 protein, Δ-CCDC hTERT-RPE1 and controls were seeded, fixed and immunostained as described previously, but only CENTRIN3 was used for the co-localisation at the centrosome.

Δ-CCDC hTERT-RPE1 and controls were proceeded further for cilia formation analysis, as described previously. Data were analysed using GraphPad Prism 6 software. Data from patient and control cells were compared using the one-way ANOVA with a post-hoc Tukey’s test. Error bars reflect the standard error of the mean (SEM).

### Animals

#### Generation of a mouse model expressing a GPATCH11 protein analogous to the human mutant variant

To express a GPATCH11 protein analogous to the mutant variant found in humans, we employed the CRISPR-Cas9 methodology to delete *Gpatch11* exon 5, which corresponds to exon 4 in humans (**Fig. 5a**). To achieve this, we designed two sgRNA guides: sgRNA_DEL1 (5’-ATACCATCTCCTAAAAAAGG-3’) and sgRNA_DEL2 (5’-TTCGGGTGATATCAATTATA-3’) from the mouse *Gpatch11* intron 4 and intron 5 sequences, respectively, using the CRISPOR software. C57BL/6J female mice (4 weeks old) were super ovulated by intraperitoneal injection of 5 IU PMSG (SYNCRO-PART® PMSG 600 UI, Ceva) followed by 5 IU hCG (Chorulon 1500 UI, Intervet) at an interval of 46–48 h and mated with C57BL/6J male mice. The next day, zygotes were collected from the oviducts and exposed to hyaluronidase (Sigma-Aldrich) to remove the cumulus cells and then placed in M2 medium (Sigma-Aldrich) into a CO2 incubator (5% CO2, 37 °C). Recombinant Cas9 protein, tracrRNA and crRNA were purchased from Integrated DNA technologies. tracrRNA:crRNA duplex (200ng/uL) and Cas9 protein (2 µM) were mixed for 10 minutes at room temperature to form an active ribonucleoprotein (RNP) complex and then added to in Opti-MEM buffer (ThermoFisher Scientific). A glass chamber equipped with platinum plate electrodes with a 1 mm gap (NEPA GENECo. Ltd) was filled with a medium (7 μL) containing RNPs. Multiple batches of zygotes were carefully aligned between the electrodes and exposed to repeated electroporation pulses using the NEPA21 electroporator, facilitating the entry of RNPs and ssODN into the zygotes. Surviving zygotes were placed in KSOM medium (Merck-Millipore) and cultured overnight until they reached the two-cell stage. Subsequently, they were transferred into the oviduct of B6CBAF1 pseudo-pregnant females. Newborn mice were genotyped using genomic and exonic DNA through PCR amplification, followed by Sanger sequencing employing appropriate primers (**Supplementary Table 2**). TIDE analysis (https://tide-calculator.nki.nl/; data not shown) was conducted. F0 founder mice carrying the deletion were bred with C57BL/6J wildtype animals to eliminate potential off-target effects. The resulting backcrossed heterozygous mice were subsequently interbred to produce homozygous mice harboring the GRCm39 chr17:1779147497-1779147632 deletion (mutant *Gpatch11Δ5/Δ5* mouse). Wildtype C57BL/6J mice served as the reference group in all analyses. Animal procedures were conducted with approval from the French Ministry of Research, in accordance with the French Animal Care and Use Committee of Paris Descartes University (APAFIS#31460), and in compliance with ethical principles within the LEAT Facility of Imagine Institute.

#### Western Blot analysis

15-day-old *Gpatch11^Δ^*^5^*^/Δ^*^5^ and wild-type mice were sacrificed by cervical dislocation and enucleated. The eye globes were dissected in PBS under a microscope to recover the retina. The tissue was lysed with RIPA-PIC (Thermo Fisher Scientific) and proceeded further for Western blot analysis as described previously, but to compare GPATCH11 abundance across samples, membranes were secondarily incubated with rabbit anti-γ-TUBULIN4B and goat anti-rabbit IgG-HRP primary and secondary antibodies, respectively. GraphPad Prism 6 software was used for statistical analyses. The abundance of GPATCH11 relative to γ-TUBULIN was estimated. The significance of variations among samples was estimated using the Student’s t test.

#### Electroretinographic analysis of mice model

*Gpatch11^Δ^*^5^*^/Δ^*^5^ and wildtype mice aged 15 days to 6 months underwent retinal function analysis via electroretinography (ERG) using the Celeris (Diagnosys LLC) apparatus. The mice were kept in darkness the day before ERG. Mice were lightly anesthetized by intramuscular injection of Ketamine (120 mg/kg; Vibrac France) and Rompun 2% xylazine (16 mg/kg; Bayer) and maintained at 37°C on a platform throughout the procedure. The animal’s pupil was dilated using Tropicamide (Mydriaticum 2 mg/0.4 mL) and phenylephrine (Néosynéphrine Faure 5%, Europhta), and a drop of sterile contact gel (LacriGel, Lacryvisc gel ophthalmic, Novartis) was applied to the eye to protect the cornea and ensure electrical contact. Electrodes were placed on the cornea, and a miniature subdermal electrode were inserted at the base of the mice’s tails for recording. To assess rod function, dark-adapted mice were exposed to light stimulations of 0.01, 0.1, 1, and 3 cd.s/m2. For cone function assessment, in light-adapted conditions mice were subjected to light stimulation of 3 and 10 cd.s/m2 (light adaptation during 8 minutes to saturate/bleach the rods). The amplitudes of the “a” and “B” waves, which measure photoreceptor and bipolar cell activation, respectively, were analyzed using Diagnosys software. Statistical analysis was conducted using GraphPad Prism 6 software, and the significance of the difference in a-wave and B-wave amplitudes between age-matched mutant *Gpatch11^Δ^*^5^*^/Δ^*^5^ and wildtype mice was determined through post hoc Sidak’s test following two-way ANOVA.

#### Longitudinal analysis of retina histology

Eyes from mutant and wildtype mice, aged 15 days to 6 months, were collected following previously described procedures. The eyes were fixed in 4% PFA overnight at 4°C. Subsequently, the specimens underwent a series of steps, including washing in PBS 1X, dehydration using an ethanol gradient, embedding in paraffin, and sectioning into 4 μm thick sections. For staining, paraffin-embedded eyes were deparaffinized using Histoclear (Electron microscopy sciences) and rehydrated by incubation in baths of decreasing ethanol concentration. Subsequently, eye sections were stained with Hematoxylin-Eosin (HE) and imaged using a NanoZoomer S210 microscope from Hamamatsu Images were analyzed using NDP view software (Hamamatsu). The thickness of the outer nuclear layer (ONL) and inner nuclear layer (INL) of the retina was measured at various distances (0.25, 0.50, 0.75, 1, 1.25, 1.50, 1.75, and 2 mm) from the optic nerve. Three mice from each group were included in the analysis. Data were analysed using GraphPad Prism 6 software. The ONL thickness of age-matched mutant *Gpatch11^Δ^*^5^*^/Δ^*^5^ and wildtype mice was compared using a two-way ANOVA with a post hoc Sidak’s test.

#### Longitudinal analysis of Gpatch11 mRNA and protein expression in the retina

For mRNA expression study, RNAScope analysis was conducted on eye sections obtained as previously described. Specifically designed RNAScope probes, manufactured by Advanced Cell Diagnostics (ACD), were employed to target *Gpatch11* mRNA. Image capture was performed using a Spinning Disk Zeiss microscope equipped with a 63x/1.4 Oil objective. For analysis, we initially utilized Fiji^37^ to prepare images for RNAScope quantification. The quantification was then carried out using Icy^40^ (v2.4.3.0) with the Spot detector plugin. Image figures were generated using FigureJ^38^. The number of *Gpatch11* spots per tissue area of age-matched mutant *Gpatch11^Δ^*^5^*^/Δ^*^5^ and wildtype mice was compared using a two-way ANOVA with a post hoc Sidak’s test. GraphPad Prism 6 software was used for statistical analyses.

For immunohistochemistry analysis of proteins deparaffined eye sections were incubated in Trisodium citrate (10 mM, pH 6) and Tween (0.05%) for 30 minutes at 95°C to achieve antigen retrieval. Subsequently, the sections were blocked for 1 hour with blocking solution containing 5% BSA in PBS. Primary antibodies were prepared in blocking solution and eye sections were further incubated overnight at 4°C in humidifying chamber. Mice eyes were immunostained using rabbit anti-GPATCH11 (1:100, Abcam) primary antibody. Sections were washed three times with PBS and incubated for 1 hour with Alexa 555-conjugated donkey anti-rabbit (IgG, 1:200, Life Technologies ThermoFisher Scientific). DAPI (Roche, Mannheim, Germany) was diluted with PBS to final 1.25 μg/mL and used to label nuclei and sections were washed with PBS and then mounted with Fluoromount medium (Sigma) under glass coverslip. Images were recorded on Spinning Disk Zeiss microscope (Zeiss) using a 63x/1.4 Oil objective. For the analysis, we first used machine learning for the detection of GPATCH11 and tissue detection using Ilastik^12^ (v1.3.3post3). Then we measured the mean intensity of GPATCH11 with a Fiji^37^ macro. Image figures were made through FigureJ^38^. The mean intensity of GPATCH11 of age-matched mutant *Gpatch11^Δ^*^5^*^/Δ^*^5^ and wildtype mice was compared using a two-way ANOVA with a post hoc Sidak’s test. Data were analysed using GraphPad Prism 6 software.

Mice eyes were also immunostained using rabbit anti-Blue S-OPSIN (1:100, Invitrogen), anti-M-OPSIN (1:1000, Invitrogen) and mouse anti-RHODOPSIN (1:500, Novus) primary antibody and Alexa 555-conjugated donkey anti-rabbit and 633-conjugated goat anti-mouse (1:200, Life Technologies ThermoFisher Scientific).

### Behavioural characterization of mice model

#### Novel object recognition paradigm (NOR)

We employed a modified version of the NOR (Novel Object Recognition) test, as previously described by reference^41^. Test sessions were recorded using an overhead camera placed above a grey plastic test arena measuring 60 × 40 × 32 cm. The light intensity within the arena was maintained consistently at approximately 20 lux. Two distinct objects were utilized: a blue ceramic pot (diameter: 6.5 cm; maximum height: 7.5 cm) and a clear glass (diameter: 8.5 cm; maximum height: 7 cm). The designation of the novel object and the placement of objects on either the left or right side of the arena were counterbalanced within each experimental group. The NOR paradigm comprised three phases conducted over three consecutive days: a habituation phase, a training phase, and a testing phase. At the outset of each exposure, mice were consistently placed in the center of the arena.

On day 1 (habituation phase), mice were allowed 5 minutes to explore the empty arena, devoid of any objects, and were then returned to their respective home cages.

On day 2 (training phase), mice were provided with a 10-minute exploration period in which they encountered two identical objects positioned symmetrically opposite to the center of the arena. Following this exploration, they were returned to their home cages.

On day 3 (testing phase), mice were given 15 minutes to explore two objects within the same arena: a familiar object and a novel one. For the purpose of scoring, exploration behaviours included sniffing, licking, nose or front leg touching of objects, or proximity (≤ 1 cm) between the mouse’s nose and the object. Immobility or being on top of an object was not considered exploration.

Preference for the novel object was calculated as the time spent exploring the new object divided by the total time spent exploring both objects. The discrimination index was computed as the difference between the time spent exploring the new object and the time spent exploring the familiar object, divided by the total time spent exploring both objects. Video recordings of the behaviour were evaluated by an observer who was blinded to the identity of the mice. Behavioural data were estimated to be statistically significant when p≤0.05 by Student’s t test. Data were analyzed using GraphPad Prism v10.0.2 software.

#### 3-foot shock contextual fear conditioning (CFC)

Individual mice were subjected to testing within conditioning chambers (Bioseb, France), each measuring 25 × 25 × 25 cm. These chambers were situated within a larger, insulated plastic cabinet (67 × 55 × 50 cm, Bioseb), providing isolation from external light and noise interference.

The chamber floors were composed of 27 stainless steel bars, wired to a shock generator equipped with a scrambler for the administration of foot shocks. Mouse movements within the chamber generated signals, which were recorded and analyzed using a high-sensitivity weight transducer system. The analysis focused on the ratio of time spent active to time spent immobile, with a particular emphasis on measuring freezing behaviour.

The contextual fear conditioning (CFC) procedure unfolded over two consecutive days. On the first day (training), mice were introduced into the conditioning chamber and subjected to three foot shocks, each lasting 1.5 seconds and delivered at intervals of 60, 120, and 180 seconds after the animals were initially placed in the chamber. Following the final shock, the mice were returned to their respective home cages, 60 seconds later.

Contextual fear memory was assessed 24 hours after conditioning (testing) by reintroducing the mice to the conditioning chamber. During this 4-minute retention test, freezing behaviour was evaluated. The assessment of freezing behaviour was conducted automatically using Packwin 2.0 software (Bioseb), with freezing defined as a period of immobility lasting at least two seconds. Behavioural data were estimated to be statistically significant when p≤0.05 by Student’s t test. Data were analyzed using GraphPad Prism v10.0.2 software.

#### Open field test (OFT)

This test leverages rodents’ inherent aversion to well-lit areas. Each mouse was placed in the center of a 43 × 43 cm chamber specifically designed for the Open Field Test (OFT) and allotted a 30-minute exploration period. Continuous monitoring of the mice during each testing session was conducted using an infrared light beam activity monitor operated by the actiMot2 Software (PhenoMaster Software, TSE). To gauge overall motor activity, we quantified the total distance traveled (ambulation). Concurrently, anxiety levels were assessed by measuring both the time spent and the distance traveled by the mice in the center of the open-field chamber compared to the periphery. Behavioural data were estimated to be statistically significant when p≤0.05 by Student’s t test. Data were analyzed using GraphPad Prism v10.0.2 software.

#### Morris Water Maze (MWM)

The Morris Water Maze (MWM) is a circular swimming pool with a white interior (diameter: 200 cm, wall height: 60 cm), situated in a room adorned with various distant visual cues. The pool was filled with water (depth: 50 cm) maintained at a constant temperature of 22°C ± 1°C. To obscure visibility, non-toxic white paint was added to the water, rendering it opaque. Submerged 1.0 cm below the water’s surface was a 12 cm round platform. The swimming pool was located in a well-lit room and featured additional cues such as geometric shapes and high-contrast patterns mounted on the walls. Each daily trial comprised four swimming attempts, during which each mouse was gently placed in the pool facing the wall, with the objective of reaching the hidden platform within 120 seconds. A trial concluded once the mouse successfully reached the platform and remained there for 5 seconds. Subsequently, the mice were returned to their home cages, with a 5-minute inter-trial interval between attempts. Different release points were utilized on a daily basis to prevent a consistent starting advantage. The time taken by the mice to reach the platform was used as a measure of their performance in locating the target. Behavioural data were estimated to be statistically significant when p≤0.05 by Student’s t test. Data were analyzed using GraphPad Prism v10.0.2 software.

#### Analysis of retina histology

Testis from mutant and wildtype mice, aged 21 days, were collected and then fixed in 4% PFA overnight at 4°C and preceeded subsequently as previously described.

#### Transcriptomic analysis of mutant and wildtype mouse retina

Total RNA was extracted from retinal tissue using the RNAeasy Mini kit, following the manufacturer’s protocol (Qiagen). The concentration and purity of the total RNA were assessed using an Agilent 2100 Bioanalyzer.

For RNA-Seq analysis, retinas from both wildtype (n=5) and *Gpatch11^Δ^*^5^*^/Δ^*^5^ (n=5) mice at 15 days were used. RNA-Seq libraries were constructed from 1 μg of total RNA using the TrueSeq Stranded mRNA Sample Prep kit (Illumina), and paired-end sequencing with 100 base pair read length was performed on a Novaseq 6000 Illumina sequencer. Pass-filtered reads were aligned to the Ensembl genome assembly GRCm39.108 using STAR 2.7.10b.

Differential expression analysis was conducted using FeatureCounts^42^ to generate a count table of gene features. For gene-level analysis, the EdgeR R package was employed for normalization, differential expression analysis, and computation of TPM (Transcripts Per Million) values^43^. Differentially expressed genes (DEGs) were filtered based on a fold change (FC) greater or <2, a False Discovery Rate (FDR) lower than 0.05, and a minimum expression of five TPM.

Alternative splicing analysis was performed using rMATS^44^, allowing the assessment of differential exon usage for various event types (*e.g.*, skipped exons, alternative 5’ and 3’ splice sites, retained introns, and mutually exclusive exons). rMATS was run with default settings using sorted BAM files produced by STAR (two-pass). Identified splicing changes were filtered based on a minimum count of 2 in at least one sample, an FDR of <0.05, and a change in inclusion-level difference of more than 20%.

A comprehensive analysis of gene lists, including enrichment in biological pathways and gene annotations from both differential expression and alternative splicing analysis, was conducted using the Gene Ontology classification system, and Metascape was utilized for this purpose^45^. Data mining and graphical representation of pathway data were performed using R packages, including GOPlot^46^. The raw data have been deposited in BioStudies and are accessible with the identifier S-BSST1157.

The downregulation of *Arr3* in *Gpatch11^Δ^*^5^*^/Δ^*^5^ retina, compared to their wildtype counterparts, was confirmed through RT-PCR using specific primers *Arr3*_forward: 5’-CAAGATTGCAGTTGTCCAGA-3’ and *Arr3*_reverse: 5’-TGGCTGGCAGACCTCCT-3’, following previously described procedures.

### Mass spectrometry (MS) analysis of mutant and wildtype mouse retina

#### TTP-Liquid Chromatography Tandem Mass Spectrometry (LC-MS/MS)

We subjected cell lysates (50 µg) to digestion using S-Trap™ micro spin columns (Protifi, Huntington, USA), following the manufacturer’s instructions. The eluted peptides were vacuum-dried and then reconstituted in a 125 µl solution containing acetonitrile (2%) and formic acid (0.1%) in HPLC-grade water before being prepared for MS analysis.

For analysis, input samples (1 µL) were injected into a nanoelute HPLC system (Bruker Daltonics, Germany) coupled to a timsTOF Pro mass spectrometer (Bruker Daltonics, Germany). High-performance liquid chromatography (HPLC) separation was performed at a flow rate of 250 nL/min using a packed emitter column (C18, 25 cm × 75 μm, 1.6 μm; Ion Optics, Australia) with a 70-minute gradient elution profile: from 2% to 13% solvent B over 41 minutes, followed by 13% to 20% over 23 minutes, then 20% to 30% over 5 minutes, 30% to 85% for 5 minutes, and finally, 85% for 5 minutes to wash the column.

Mass-spectrometric data were acquired using the parallel accumulation serial fragmentation (PASEF) acquisition method in data-dependent acquisition (DDA) mode. Measurements were conducted within the m/z range of 100 to 1700 Th (Thomson). The ion mobility values ranged from 0.75 to 1.25 V s/cm² (1/k0). The total cycle time was set to 1.17 seconds, and 10 PASEF MS/MS scans were performed.

#### Data processing LC-MS/MS acquisition

The mass spectrometry (MS) data files were processed using MaxQuant software version 2.1.3.0 and searched against the Mus musculus database from SwissProt and TrEMBL (release 11/2021, containing 88,132 entries) using the Andromeda search engine. We configured the search with an initial mass deviation of 4.5 ppm for parent masses and 20 ppm for fragment ions. The minimum peptide length was set to 7 amino acids, and we enforced strict specificity for trypsin cleavage, allowing up to two missed cleavage sites.

Carbamidomethylation (Cys) was specified as a fixed modification, while oxidation (Met) and N-terminal acetylation were set as variable modifications. We also enabled ‘Match between runs’. For label-free quantification (LFQ), the minimum ratio count was set to 1. False discovery rates (FDRs) were controlled at both the protein and peptide levels, set to 1%. Scores were calculated within MaxQuant as previously described^47^.

To ensure data quality, we removed reverse and common contaminant hits from the MaxQuant output. Protein quantification was achieved using the MaxQuant label-free algorithm with LFQ intensities^47,48^. We analyzed four independent lysate replicates for both wildtype and homozygote *Gpatch11*^Δ5/Δ5^ mutant samples using Perseus software (version 1.6.15.0), which is freely available at www.perseus-framework.org^49^.

For statistical comparisons, we divided the data into two groups, each consisting of up to 4 biological replicates. We filtered the data to retain only proteins with at least 3 valid values in at least one group. Missing data points were imputed by generating a Gaussian distribution of random numbers with a standard deviation of 33% relative to the standard deviation of the measured values, along with a 1.8 standard deviation downshift of the mean to simulate the distribution of low signal values.

Statistical analysis was performed using a t-test with an FDR threshold of <0.05 and S0=0.01. Volcano plots of proteins were generated in Perseus using logarithmized LFQ intensities with an FDR threshold of <0.05 and S0=0.01.

The raw data from the lysates have been deposited in ProteomeXchange via the PRIDE database and can be accessed with the identifier PXD041849.

## Data Availability

All data produced in the present work are contained in the manuscript

## ACKNOWLEDGMENTS

This work was supported by grants from Retina France; Fondation JED Belgique; Fondation Visio to JMR and IP. Fondation des Aveugles de France, PhD international Institut Imagine to AZ and by grant # 310030_204285s from the Swiss National Science Foundation to CR. JMR is member of the European Reference Network for Rare Eye Diseases (ERN-EYE), which is co-funded by the Health Program of the European Union under the Framework Partnership Agreement n°739534. We thank the Transgenesis, Neurobehavioural and metabolism, Necker Bioimage Analysis, Cell imaging, Histology, Cytometry, Genomic and Proteomic platforms for valuable discussions and help. A special thank for Nicolas Cagnard for the submission to BioStudies. The authors would like to acknowledge all patients and their families for participating in this study. The authors are also grateful to Virginie G. Peter and Raquel Rodriguez for their help throughout the study.

## AUTHOR CONTRIBUTIONS

A.Z. performed *in vitro and in vivo* experiments, analyzed the data and wrote the paper

P.D., L.F-T. performed creation of animal models

J.A C.G I.A. M.R. N.B. AL. B. N.P-S. D.A. J.K. C.S. L.C.S. M.S. A.B.S. collected clinical data

S.M. performed behavioural analysis of mice

N.G. created analytic tools

J.R analyzed data from RNASeq

C.G analyzed data from mass spectrometry

S.B. collected skin biopsy from patients

I.P analyzed exome data from Family 1

J.A analyzed exome data from Family 2

C.T. analyzed exome data from Family 3

K.K. M.Q. C.R. analyzed exome data from Family 4

J-M.R. I.P. supervised the research, designed the experiments and wrote the paper

All authors discussed the results and participated in manuscript preparation and editing

