## Supplementary Information for "*GPATCH11* variants cause mis-splicing and early-onset retinal dystrophy with neurological impairment"

**Supplementary Table 1: Founder effect in the three families carrying the c.328+1G>T (p.?) *GPATCH11* variant.** Chr.: Chromosome, RS: Reference Single nucleotide, REF: nucleotide of reference, ALT: alternative nucleotide, AF: allelic frequency**.**

| Chr. | Position | RSL | REF | ALT | AF (alt) | F1:III-6 | F1:IV-1 | F1:IV-2 | F2:II-2 | F3:II-2 |
|---|---|---|---|---|---|---|---|---|---|---|
| 2 | 36356628 | rs113549769 | G | A | 0.094% | AA | GA | AA | GA | GA |
| | 36548827 | | G | C | 0.00% | GG | GC | GG | GG | GG |
| | 36755004 | | G | T | 0.00% | **GG** | **GG** | **GG** | GT | **GG** |
| | 36878185 | | T | C | 0.793% | **TT** | **TT** | **TT** | TC | **TT** |
| | 36982026 | rs1251060661 | T | TA | 0.076% | **TT** | **TT** | **TT** | TTA | **TT** |
| | 37083479 | | A | C | 0.00% | **AA** | **AA** | **AA** | AA | **AA** |
| | 37090723 | rs1407579456 | G | T | 0.00% | **TT** | **TT** | **TT** | TG | **TT** |
| | 37092169 | rs574837250 | C | T | 0.00% | **CC** | **CC** | **CC** | CT | **CC** |
| | 37171345 | rs11569760 | C | G | 0.678% | **CC** | **CC** | **CC** | CG | **CC** |
| | 37372210 | rs113488761 | T | C | 0.541% | **TT** | **TT** | **TT** | TC | **TT** |
| | 37817975 | rs59076457 | G | A | 2.00% | AA | AA | AA | GG | GG |
| | 38017550 | | | | | GG | GG | GG | TT | TT |

**Supplementary Table 2: Sequences and positions of PCR and RT-PCR primers specific to human *GPATCH11* and mouse *Gpatch11* sequences, respectively.**

| Targeted region | Orientation | Position | Sequence (5' > 3') |
|---|---|---|---|
| *GPATCH11* DNA exon 3 | *Forward* | Intron 3 | TTATGCCATTGCATTACAGC |
| | *Reverse* | Intron 4 | GATTAAAGCATCAGCTTTGG |
| *GPATCH11* DNA exon 4 | *Forward* | Intron 4 | GCATTGTAAAGTTGCATACT |
| | *Reverse* | Intron 5 | TGAACTGTAACAGTATTATC |
| *GPATCH11* DNA exon 5-6 | *Forward* | Intron 5 | CCCTAGTGTTTTCTACTATTAGTG |
| | *Reverse* | Intron 7 | GGGAAATCACAGGGGGTGCC |
| *GPATCH11* cDNA exon 3-4-5 | *Forward* | Exon 2/3 | GTCCAAGAAGATATCAGACC |
| | *Reverse* | Exon 5/6 | AGTCGCATTCGAAACTGTTC |
| *GPATCH11* cDNA exon 3-4-5-6 | *Forward* | Exon 2/3 | GTCCAAGAAGATATCAGACC |
| | *Reverse* | Exon 7 | CAACCAGTACCATGCTTCC |
| *Gpatch11* DNA exon 5 | *Forward* | Intron 5 | CCAGAATTCAGTTTACAGTT |
| | *Reverse* | Intron 6 | CCATTTCATCAAAATTTACTTG |
| *Gpatch11* cDNA exon 4-5-6-7 | *Forward* | Exon 3 | CATTCATGTTCAAGAAGATGTC |
| | *Reverse* | Exon 8 | GAATATTCTTCTGGGCATCC |

### Supplementary Table 3: Splicing events of 12 DEG and DSG genes.

| Gene.name | event | RNA | deltaPSI | KI_meanPSI | WT_meanPSI | KI.1_IJC | KI.2_IJC | KI.3_IJC | KI.4_IJC | KI.5_IJC | KI.1_SJC | KI.2_SJC | KI.3_SJC | KI.4_SJC | KI.5_SJC | WT.1_IJC | WT.2_IJC | WT.3_IJC | WT.4_IJC | WT.5_IJC | WT.1_SJC | WT.2_SJC | WT.3_SJC | WT.4_SJC | WT.5_SJC |
|---|---|---|---|---|---|---|---|---|---|---|---|---|---|---|---|---|---|---|---|---|---|---|---|---|---|
| *Arr3* | SE | in frame | -0,459 | 0,34 | 0,8 | 31 | 47 | 25 | 10 | 7 | 45 | 57 | 34 | 18 | 15 | 84 | 102 | 77 | 60 | 73 | 10 | 16 | 28 | 17 | 9 |
| *Asap3* | RI | frameshift | 0,211 | 0,52 | 0,31 | 60 | 42 | 26 | 45 | 40 | 15 | 24 | 10 | 14 | 10 | 6 | 26 | 12 | 11 | 10 | 13 | 19 | 6 | 9 | 7 |
| *Cabp4* | A3SS | frameshift | 0,3 | 0,61 | 0,32 | 133 | 128 | 56 | 54 | 92 | 21 | 27 | 15 | 16 | 17 | 69 | 70 | 59 | 41 | 37 | 43 | 57 | 23 | 54 | 43 |
| *Ccnl2* | SE | in frame | 0,325 | 0,57 | 0,25 | 50 | 67 | 31 | 30 | 19 | 2 | 6 | 5 | 4 | 8 | 48 | 37 | 35 | 22 | 19 | 25 | 28 | 10 | 10 | 21 |
| *Dhrs3* | SE | in frame | -0,249 | 0,43 | 0,68 | 36 | 52 | 31 | 46 | 26 | 9 | 5 | 6 | 28 | 21 | 42 | 46 | 60 | 42 | 39 | 3 | 7 | 5 | 8 | 2 |
| *Dhrs3* | A3SS | in frame | 0,204 | 0,46 | 0,26 | 147 | 173 | 111 | 134 | 113 | 63 | 61 | 83 | 48 | 57 | 58 | 164 | 122 | 96 | 139 | 144 | 138 | 194 | 100 | 96 |
| *Lbhd1* | MXE | in frame | -0,283 | 0,45 | 0,74 | 249 | 199 | 313 | 267 | 237 | 324 | 313 | 322 | 127 | 210 | 172 | 468 | 386 | 430 | 536 | 120 | 106 | 125 | 85 | 79 |
| *Lbhd1* | MXE | frameshift | -0,279 | 0,47 | 0,75 | 266 | 205 | 321 | 275 | 249 | 324 | 313 | 322 | 127 | 210 | 190 | 477 | 403 | 440 | 541 | 120 | 106 | 125 | 85 | 79 |
| *Lbhd1* | MXE | frameshift | -0,274 | 0,49 | 0,77 | 249 | 209 | 318 | 270 | 239 | 324 | 313 | 322 | 127 | 210 | 186 | 473 | 386 | 438 | 543 | 120 | 106 | 125 | 85 | 79 |
| *Lbhd1* | MXE | in frame | -0,23 | 0,55 | 0,78 | 318 | 266 | 399 | 339 | 313 | 324 | 313 | 322 | 127 | 210 | 196 | 520 | 425 | 491 | 579 | 120 | 106 | 125 | 85 | 79 |
| *Mpp4* | SE | in frame | -0,3 | 0,4 | 0,7 | 141 | 144 | 136 | 91 | 63 | 165 | 187 | 113 | 103 | 82 | 136 | 232 | 190 | 141 | 141 | 19 | 91 | 78 | 46 | 56 |
| *Pex5l* | A3SS | in frame | -0,32 | 0,39 | 0,71 | 11 | 20 | 20 | 19 | 11 | 13 | 41 | 36 | 28 | 15 | 13 | 17 | 14 | 13 | 17 | 6 | 11 | 4 | 6 | 4 |
| *Pitpnm3* | SE | frameshift | 0,244 | 0,46 | 0,21 | 27 | 16 | 13 | 12 | 12 | 24 | 41 | 16 | 10 | 2 | 4 | 19 | 15 | 6 | 12 | 9 | 77 | 38 | 13 | 30 |
| *Tulp1* | A3SS | frameshift | 0,231 | 0,61 | 0,38 | 257 | 388 | 165 | 187 | 104 | 90 | 86 | 50 | 51 | 66 | 158 | 292 | 252 | 131 | 159 | 198 | 195 | 149 | 130 | 161 |
| *Tulp1* | A3SS | frameshift | 0,233 | 0,6 | 0,37 | 250 | 368 | 157 | 181 | 100 | 90 | 86 | 50 | 51 | 66 | 147 | 279 | 238 | 125 | 157 | 198 | 195 | 149 | 130 | 161 |
| *Unc13b* | SE | in frame | -0,231 | 0,3 | 0,53 | 30 | 42 | 15 | 12 | 7 | 57 | 83 | 23 | 29 | 18 | 48 | 20 | 32 | 33 | 32 | 28 | 28 | 12 | 23 | 39 |
| *Vtn* | A3SS | frameshift | -0,276 | 0,31 | 0,58 | 84 | 124 | 84 | 61 | 71 | 25 | 52 | 74 | 50 | 59 | 81 | 107 | 106 | 80 | 82 | 7 | 16 | 24 | 23 | 18 |
| *Vtn* | A3SS | frameshift | -0,273 | 0,32 | 0,59 | 84 | 127 | 87 | 64 | 81 | 25 | 52 | 74 | 50 | 59 | 89 | 109 | 109 | 80 | 83 | 7 | 16 | 24 | 23 | 18 |
| *Vtn* | A3SS | frameshift | -0,272 | 0,31 | 0,58 | 92 | 126 | 88 | 62 | 71 | 25 | 52 | 74 | 50 | 59 | 81 | 108 | 109 | 85 | 82 | 7 | 16 | 24 | 23 | 18 |

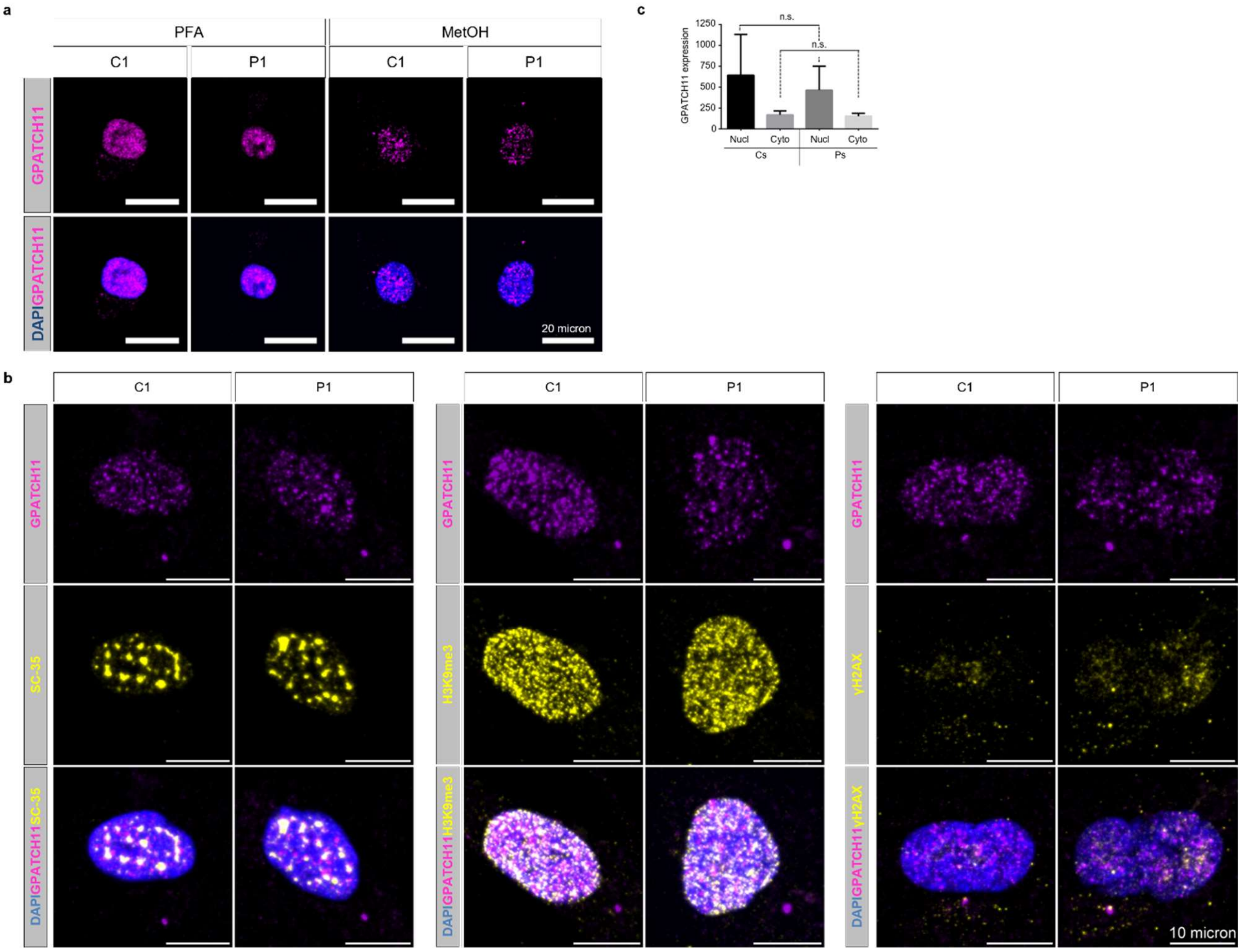


**Supplementary Fig. 1: Immunocytochemistry analysis of GPATCH11 subcellular localisation. (a)** Immunostaining of GPATCH11 (magenta) in the fibroblasts from a control (C1) and affected individuals carrying the c.328+1G>T variant in homozygosity (P1), through two different fixation methods: PFA and Methanol (MetOH). DAPI is used to label the nucleus (blue). Scale bar, 10µm. **(b)** Quantification of the cytoplasmic and nuclear GPATCH11 protein in fibroblasts of controls (Cs) and affected individuals (Ps). Graphic bars represent the mean±SEM derived from three experimental replicates. n.s., not significant. **(c)** Immunostaining of GPATCH11 (magenta), SC-35, H3K9me3 and γH2AX (yellow) in the fibroblasts from a control (C1) and affected individuals P1. DAPI is used to label the nucleus (blue). Scale bar, 10µm.

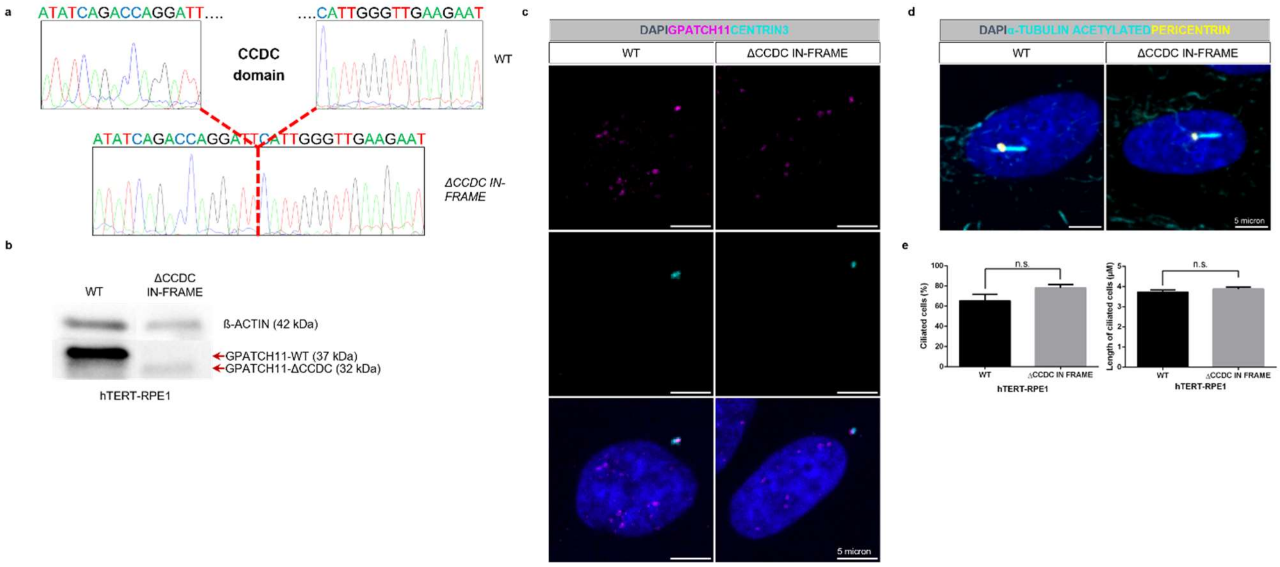


**Supplementary Fig. 2**: **Creation and ciliation analysis of Δ-CCDC hTERT-RPE1 and controls. (a)** Representative chromatogram of cDNA of the CCDC-domain deletion. **(b)** Western blot analysis of protein extracts using GPATCH11 antibody in control (WT) and Δ-CCDC hTERT-RPE1. **(c)** Immunostaining of CENTRIN3 (cyan) and GPATCH11 (magenta) proteins in control and Δ-CCDC hTERT-RPE1. DAPI is used to label the nucleus (blue). Scale bar, 5µm. **(d)** Immunostaining of ACETYLATED α-TUBULIN (cyan) and PERICENTRIN (yellow) proteins in control and Δ-CCDC hTERT-RPE1, after 24 hours of serum-free culture to promote ciliation. DAPI is used to label the nucleus (blue). Scale bar, 5µm. **(e)** Quantification of ciliated cells and length of cilia axoneme in control and Δ-CCDC hTERT-RPE1, according to immunostaining of (**d**). A minimum of 100 ciliated cells were considered for each line. Graphic bars represent the mean±SEM derived from four experimental replicates. n.s., not significant.

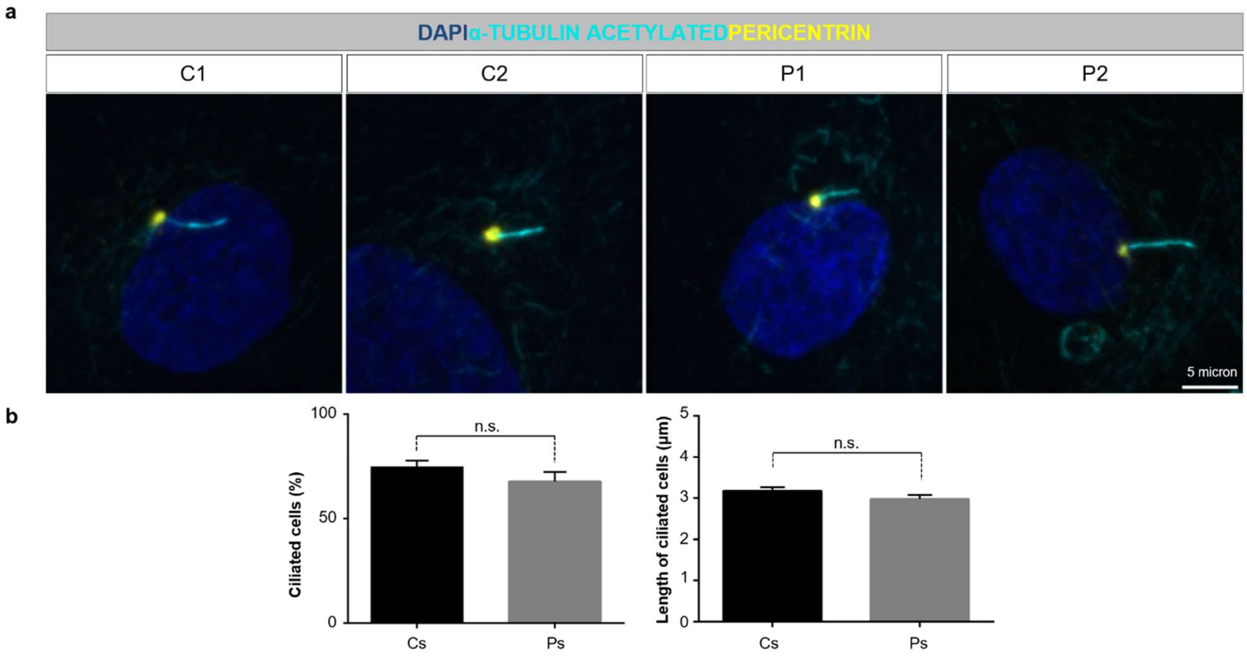


**Supplementary Fig. 3: Ciliation analysis in fibroblasts from controls and affected individuals. (a)** Immunostaining of ACETYLATED α-TUBULIN (cyan) and PERICENTRIN (yellow) proteins in fibroblasts from controls (C1, C2) and P1 and P2 affected individuals (P1, P2) after 24 hours of serum-free culture. DAPI is used to label the nucleus (blue). Scale bar, 5µm. **(b)** Quantification of ciliated cells and length of cilia axoneme in controls (Cs) and affected fibroblasts (Ps), according to immunostaining of **(a)**. A minimum of 100 ciliated cells were considered for each line. Graphic bars represent the mean±SEM derived from three experimental replicates. n.s., not significant.

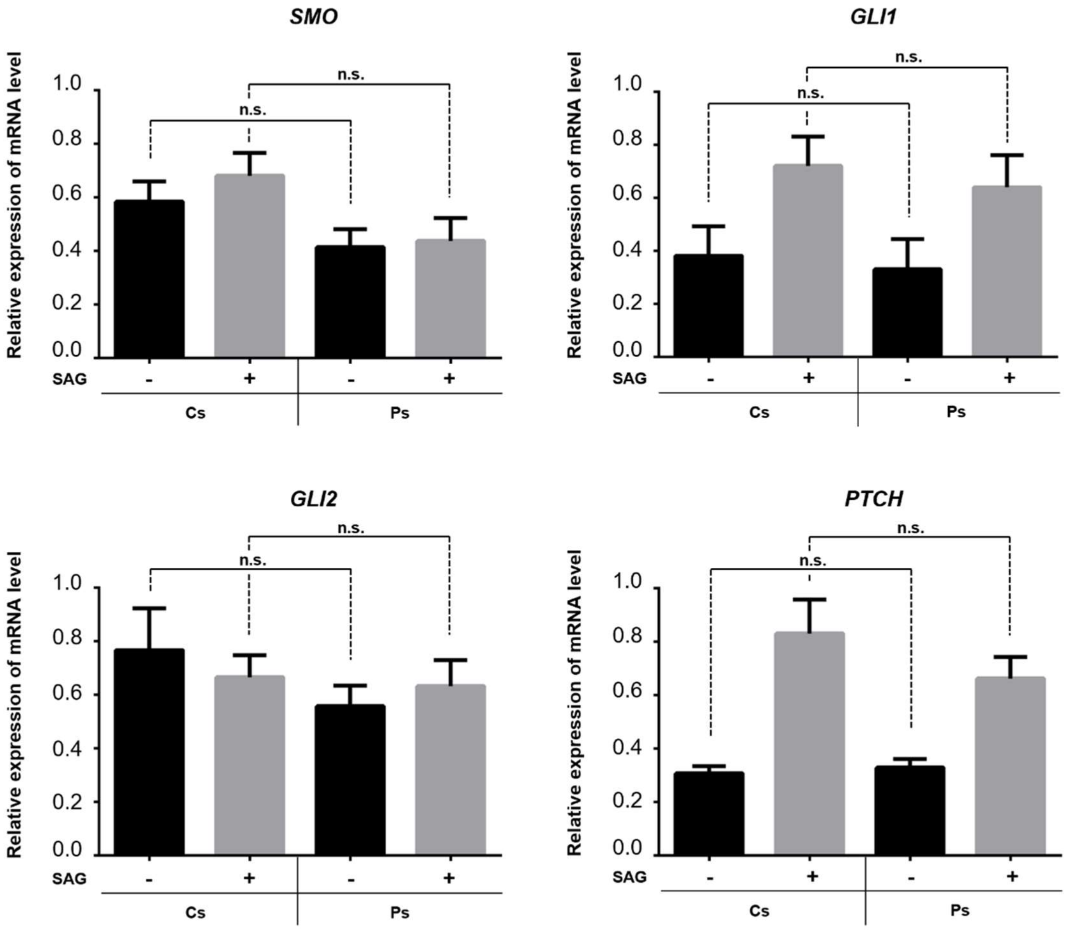


**Supplementary Fig. 4**: **SHH pathway analysis.** RT-qPCR analysis of *SMO*, *GLI1*, *GLI2* and *PTCH1* mRNAs abundance expressed in fibroblasts from controls (Cs) and P1 and P2 affected individuals (Ps) starved for 48h and untreated or treated for 24h with SAG. Graphic bars show the mean±SEM from three independent experiments. n.s., not significant.

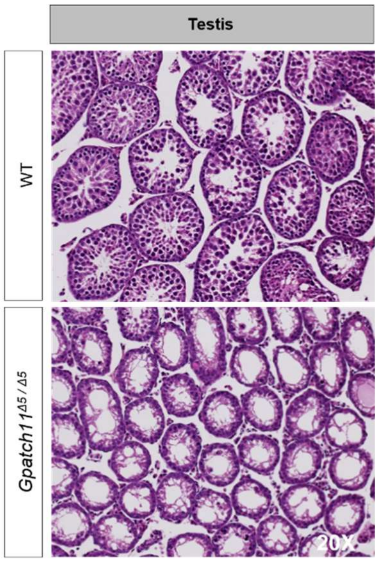


**Supplementary Fig. 5: Study of sterility on mouse model.** Histology sections of testis of *Gpatch11*$^{Δ5/Δ5}$ of 21 days compared to wildtype (WT).

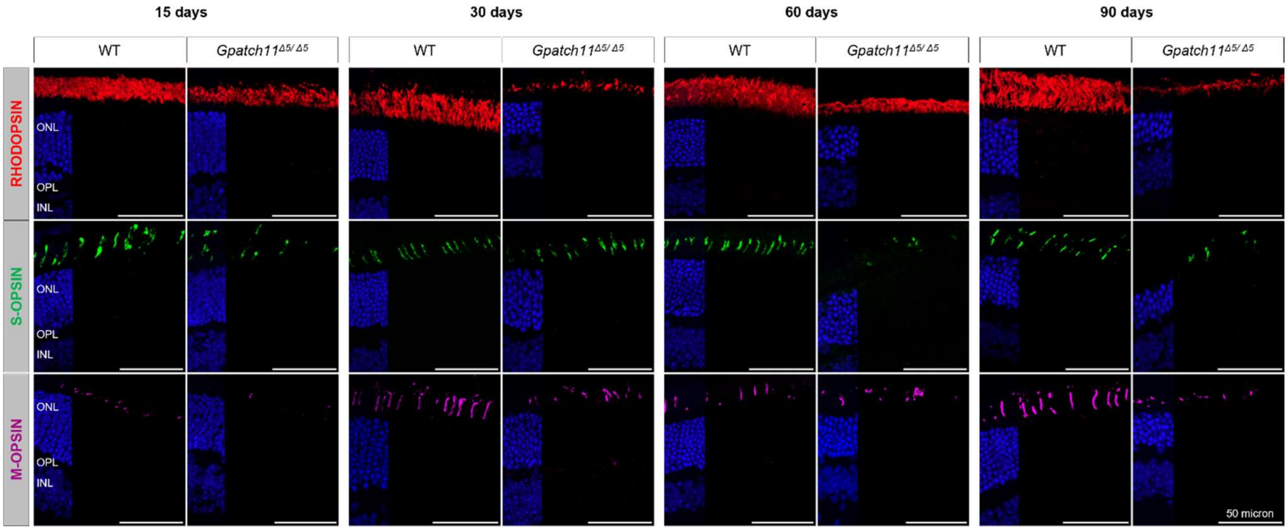


**Supplementary Fig. 6**: **Immunohistochemistry of S-OPSIN, M-OPSIN and RHODOPSIN on murine retina sections.** Immunostaining of retina sections of wildtype (WT) and *Gpatch11*$^{\Delta 5/\Delta 5}$ from 15 days to 30 days, with anti-S-OPSIN (green), anti-M-OPSIN (purple) and anti-RHODOPSIN (red) antibodies. DAPI is used to label the cell nucleus (blue). Scale bars, 50 µm.

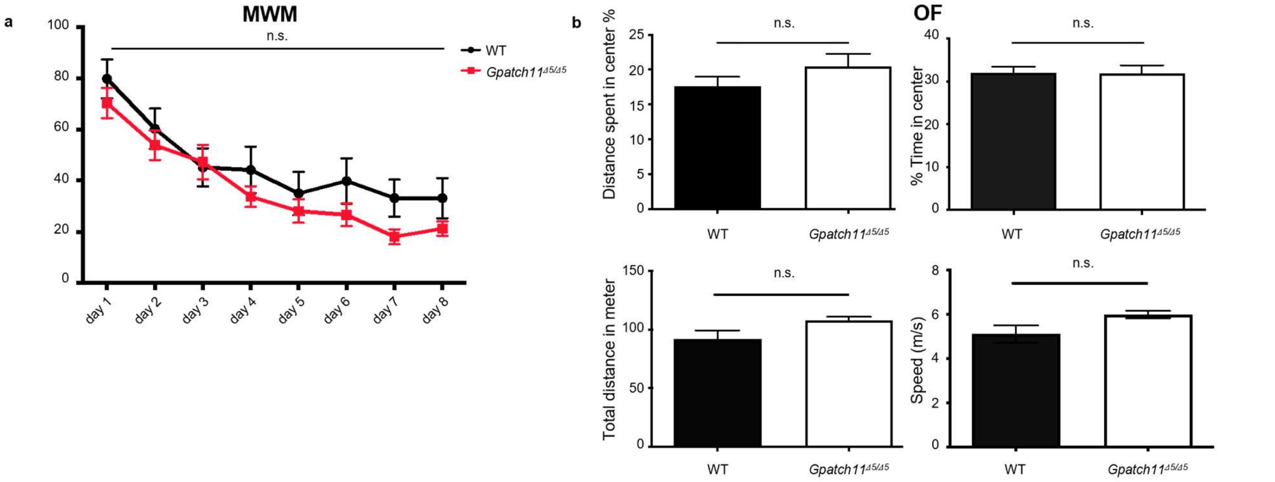


**Supplementary Fig. 7: Study of behaviour in 1-month-old wildtype (WT) and *Gpatch11*$^{\Delta 5/\Delta 5}$ mouse model. (a)** Anxiety-like behaviour assessment based on the Morris Water Maze (MWM). **(b)** Spatial memory assessment based on the Open Field (OF). Percentage of distance spent in centre, total distance in meters, percentage of time in centre and the speed were measured. Bars represent mean±SEM from 20 mice per genotype. n.s., not significant.

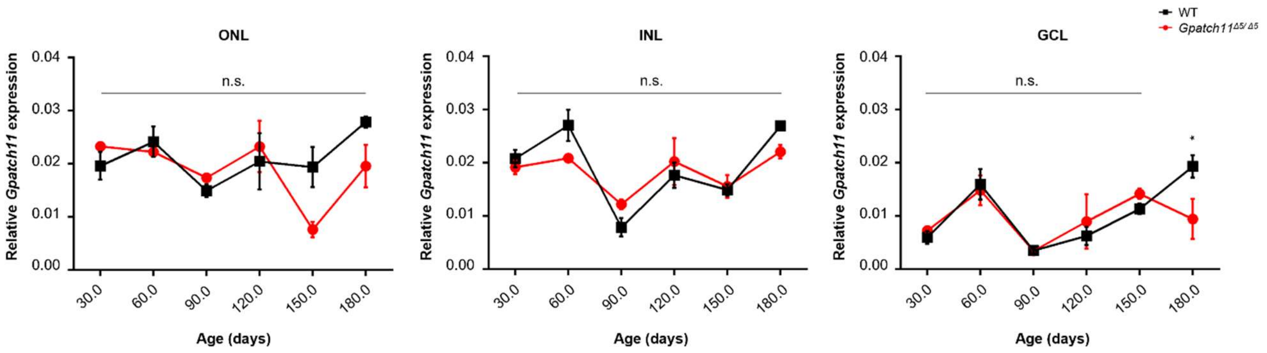


**Supplementary Fig. 8: Quantification of RNAScope analysis on murine retina sections.** Quantification of Gpatch11 transcript expression in the ONL, INL and GCL through RNAScope analysis in wildtype (WT, black) and *Gpatch11*$^{\Delta5/\Delta5}$ (red) murine retinas, from 30 to 180 days. Graphic bars represent the mean±SEM derived from three biological replicates. n.s., not significant.
